# Performance of general-population breast cancer risk prediction models in an international consortium

**DOI:** 10.64898/2026.08.20.26360899

**Authors:** Kristen D. Brantley, Thomas U. Ahearn, Emily L. Norton, Robert MacInnis, Julie R. Palmer, Renée T. Fortner, Celine M. Vachon, Laura Beane-Freeman, Amy Berrington de Gonzalez, Reuben Frost, Kimberly A. Bertrand, Gary Zirpoili, Marian L. Neuhouser, Matthew Barnett, Lauren R. Teras, James M. Hodge, Alpa V. Patel, Clara Bodelon, James V. Lacey, Emma S. Spielfogel, Thomas E. Rohan, Victoria A. Kirsh, Hilde Langseth, Kaitlyn M. Tsuruda, Roger L. Milne, Christopher Haiman, Christopher G. Scott, A. Heather Eliassen, Bernard Rosner, Walter C. Willett, Andrea Romanos-Nanclares, Yu Chen, Fen Wu, Wei Zheng, Jirong Long, Katie M. O’Brien, Dale P. Sandler, Cari M. Kitahara, Martha S. Linet, Garnet Anderson, Joseph C. Larson, I-Min Lee, Montserrat Garcia-Closas, Nilanjan Chatterjee, Peter Kraft

## Abstract

**Background:** Several breast cancer (BC) risk prediction models have been developed to provide personal risk assessments. Though individually validated, their performance has not been systematically evaluated across a wide range of populations or ages.

**Methods:** We harmonized individual-level baseline questionnaire data and incident BC diagnoses from 21 cohorts from North America, Europe, and Australia participating in the Breast Cancer Risk Prediction Project. Five-year absolute risk of invasive BC was estimated for five established risk prediction models using classical risk factors only. Discrimination was evaluated by area under the curve (AUC). Calibration was assessed using average and risk-decile specific expected to observed (E/O) ratios. Performance metrics were meta-analyzed across cohorts and models. Metaregression tested associations between cohort characteristics and performance metrics.

**Results:** This analysis included 1,595,977 women aged 20-75 years, enrolled in studies between 1976-2015, with 19,062 (1.2%) invasive BC cases ascertained within 5 years from exposure assessment. Age-adjusted AUCs were similar across models and cohorts (pooled AUCs by model: 0.57-0.58), while E/O ratios varied substantially (pooled E/O ratios by model: 0.83-1.25). Overestimation was common among predicted high-risk individuals (>3%). No appreciable differences in model performance by cohort age, birth year, race, and variable missingness emerged. Calibration improved after assigning race-specific incidence rates.

**Conclusion:** Existing BC risk prediction models provided similar risk discrimination across multiple cohorts, although there was overestimation of risk for high-risk individuals. Performance variation across cohorts was not driven by specific characteristics, which supports development of a unified risk model for diverse populations that leverages appropriate incidence rates.

**Key messages:**

- When using classical risk factor components of existing risk prediction models, we found similar discriminatory ability of models across diverse cohorts.
- Aside from underlying cancer incidence rate, which heavily influenced calibration, no cohort-specific characteristics were consistently associated with model performance.
- Risk was underestimated at lower predicted risk deciles and overestimated at higher predicted risk deciles, indicating a need to improve model fit by integrating more complex risk-factor relationships.

## INTRODUCTION

Globally, approximately 2.3 million incident breast cancer (BC) cases were recorded in 2022.^1^ Despite mortality reduction via advances in screening and treatment modalities, BC remains the second leading cause of cancer-related mortality.^1^ Risk stratification could reduce the burden of disease by identifying individuals who may benefit from chemoprevention and by enhancing screening regimens to promote early detection.^2^

Several risk prediction models estimate individualized risk of BC over specified time periods including 5-year, 10-year, and lifetime risk.^3^ Commonly implemented models include the National Cancer Institute’s BC Risk Assessment Tool (BCRAT), based on the Gail model,^4–8^ the Tyrer-Cuzick (TC) model (a.k.a. the International Breast Intervention Study (IBIS) tool),^9^ and the Breast and Ovarian Analysis of Disease Incidence and Carrier Estimation Algorithm (BOADICEA) (included in the CanRisk tool).^8,10^ More recent models include a literature-derived model using the Individualized Coherent Absolute Risk Estimation tool (iCARE-Lit),^11–13^ and the Black Women’s Health Study (BWHS) BC Risk Calculator (hereafter referred to as “BWHS RC”), which was built specifically to predict risk in Black women.^14^

Well-established BC risk factors include family history of breast or ovarian cancer, personal history of benign breast disease (BBD) or hyperplasia, menopausal status, reproductive history, hormone use, alcohol use, and BMI.^15^ While all models mentioned above consider the contribution of these classical risk factors in some way, they differ in the set of risk factors used and the relative risk contributions of these factors. Differing risk estimates can emerge from model to model, increasing the difficulty in choosing the best option for a given individual.

Moreover, these models were developed using different approaches, including direct model fitting in different, often relatively small, populations (BCRAT, BWHS) or “synthetic” models which assign and weight relative risks based on evidence from multiple studies (iCARE-Lit, TC, BOADICEA). Not all models allow for updated average cancer incidence rates and competing mortality in the target population to estimate absolute risk. To date, comparison across existing models has not been systematically performed on a large set of harmonized risk factor data across diverse populations. Here we performed comparative validation of the classical risk factor component of five BC risk prediction models in a large consortium of over 1.5 million women of diverse backgrounds.

## METHODS

### BCRPP data harmonization

The BC Risk Prediction Project (BCRPP) was created to enable large-scale comparative model validation and to develop a comprehensive BC risk prediction model with robust application across racial and ethnic groups. In 2021, BCRPP began requesting data from prospective cohorts that were designed to capture information on BC outcomes. To date, 26 cohorts have agreed to contribute to BCRPP in model development. Data governance protocols and storage infrastructure were developed to facilitate responsible data sharing with researchers developing or validating BC risk models or studying BC epidemiology generally (see **Data Availability**).

A total of 21 studies (19 full cohorts and 2 case-cohort studies) were included in this analysis, representing the U.S. (N=17), Europe (N=2), Canada (N=1), and Australia (N=1). Details on cohorts, data collection, and harmonization are in **Supplemental Methods**.

### Absolute Risk Assignment

After harmonizing data across cohorts and excluding prevalent *in situ* or invasive BC cases, 5-year absolute risk estimates for invasive BC were calculated using data for classical risk factors from five validated models: BCRAT^4–6^, iCARE-Lit^11,12^, TC (v8b)^16^, BOADICEA (v4)^10,17^, and the BWHS RC^14^. The risk period began 1 year after baseline entry. Risk was estimated using each tool as applied in practice, restricted to questionnaire-based risk factors (**Tables 1 & S1**). For iCARE-Lit, reference populations were created based on cohort demographics and incidence rates from national registries and country-specific mortality rates were applied where possible. Several model assumptions were required for estimation using the TC and BOADICEA models, as detailed pedigrees were not explicitly requested in data harmonization (**Supplemental Methods)**. To align with prospective cohort studies, risk in case-cohort studies was calculated among all subcohort members regardless of case status, and for all cases that occurred within the first five years of baseline, with sampling weights applied. To test the influence of potential uncaptured prevalent cases, risk calculations were repeated starting two years after baseline. Main analyses estimated risk for individuals aged 20-75 years via BCRAT, iCARE-Lit, TC, and BOADICEA.

To provide practical comparison of the BWHS RC to other models, we estimated 5-year absolute risk among the subset of Black women aged ≥30-70 years, using incidence and mortality rates from non-Hispanic Black women if possible (see **Table S1**).

Medians and inter-quartile ranges (IQR) were calculated by cohort and by model for risks calculated within 1-5 y and within 2-6 y from baseline.

### Model Validation

We applied a modified version of the ModelValidation function from the iCARE package in R (v.1.38.0, <u>Bioconductor download</u>) (modifications in **Supplemental Methods**), to estimate area under the curve (AUC) and 95% confidence intervals (CI) for the absolute risk within each cohort and model. To further adjust for age effects, we estimated absolute-risk based AUC within cohort-specific age quintiles and applied inverse variance weighting to determine the summary age-adjusted AUC. The ratios of the expected to observed (E/O) absolute risks were estimated overall and within deciles of expected absolute risk for each cohort. We also examined E/O ratios among individuals who had ≥3% predicted 5-year absolute risk, set as the threshold for “high-risk” by the US Preventive Services Task Force in 2019 to guide use of risk-reducing medications.^18^ Sampling weights were applied for AUC and E/O calculations in the two case-cohort studies, with adjustment made for standard errors. Decile-specific observed absolute risks and 95% CIs were plotted against expected absolute risks for each model and cohort. Random effects meta-analysis using the restricted maximum-likelihood estimator with inverse variance weighting summarized AUCs and E/O ratios for each model across cohorts.

We tested the influence of cohort characteristics such as average age, birth year, race, and risk factor missingness on the performance of each model by meta-regressing cohorts’ transformed age-adjusted AUCs and E/Os on these characteristics. We used the transformations log(qnorm(AUC)) and log(|E/O-1|) to better approximate a normal distribution in the dependent variables. Slopes, 95% confidence intervals, and p-values were estimated using the metafor package in R. Model performance was tested separately among Black women in data from cohorts with ≥100 invasive BCs among Black women.

To better mimic the ideal scenario, we visualized model calibration among those cohorts with no model-specific risk factors missing by design (**Figure 1**, **Table 1**).

**Figure 1.**
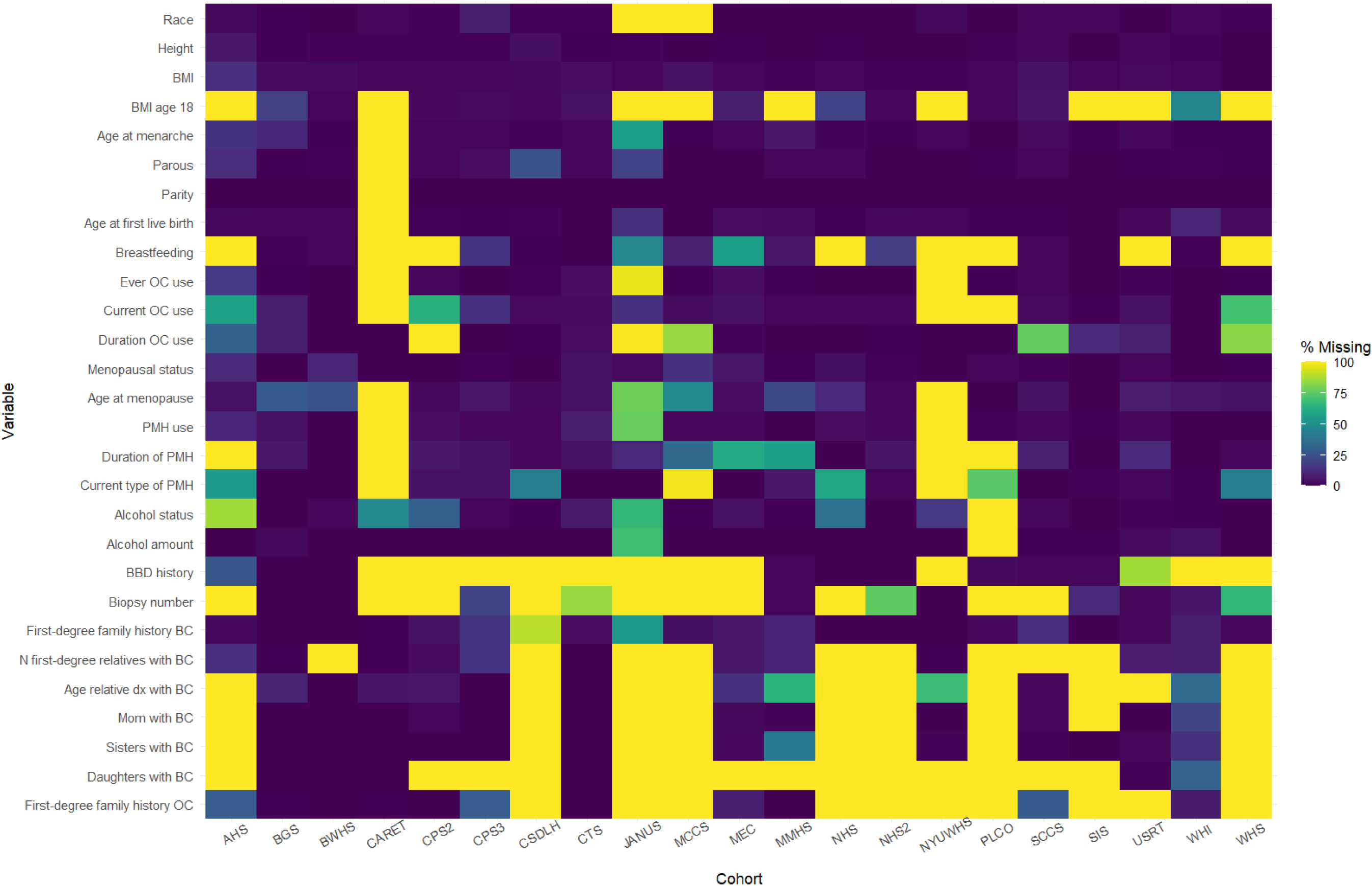
Percentage of risk factor missingness by cohort in BCRPP. Colors indicate percent of cohort missing the variable of interest. 100% missingness (i.e., data not available/provided for BCRPP) is designated by yellow, while dark blue represents 0% missing. Risk factors listed are those used in any of the models compared (BCRAT, BOADICEA, iCARE-Lit, Tyrer-Cuzick, BWHS risk calculator, see Table S1) and requested as part of the BCRPP harmonized data dictionary. Missingness counts only eligible individuals, e.g., % missing age at first birth is calculated among parous individuals only. If 100% missing for the main variable (e.g., parous), all related variables (age at first live birth, parity) are set to 100% missing. Logic follows for current OC use (% among premenopausal women only), alcohol amount (% among current drinkers), age at menopause (% among those postmenopausal), and PMH duration (% among postmenopausal women who report ever using PMH). BBD: benign breast disease (personal history of); PMH: postmenopausal hormone therapy; OC: oral contraceptive

**Table 1.** Features of existing BC risk prediction models.

| Feature | BCRAT (Gail) | IBIS (Tyrer-Cuzick) | BOADICEA (CanRisk) | BWHS BC Risk Calculator | iCARE-Lit |
| --- | --- | --- | --- | --- | --- |
| Target population | General population | General and familial/high-risk populations | General and familial/high-risk populations | General population | General population |
| Source of relative risk | Logistic regression on case-control data | Synthetic using published data | Segregation analysis on UK family data, and synthetic using published data | Logistic regression on case-control data | Synthetic using published data |
| Source of age-specific incidence rates | US race specific SEER rates from 1980's-90's | U.K. cancer registry, 2005-2009 | Country-specific registries: Australia, Canada, Denmark, Estonia, Finland, France, Iceland, Netherlands, New Zealand, Norway, Slovenia, Spain, Sweden, UK, USA | US SEER 2000-2016, non-Hispanic Black women | US SEER 2000-2009; UK National Disease Registration Service (NDRS) 2005 |
| Incorporation of race/ethnicity | Separate RR coefficients and incidence rates for White, Black, and Asian/PI women | Not incorporated | Not incorporated | RR coefficients and incidence rates for Black women | Flexible assignment of incidence rates based on cohort distribution |
| Risk factors applied in this analysis | Age, menarche, age at first birth, no. 1st-degree relatives with BC, no. biopsies, race/ethnicity | Age, menarche, parity, age at first birth, menopause, BMI, height, HRT, family history (1st degree), | Age, menarche, parity, HRT, BMI, alcohol, family history (1st degree), and residual polygenic component | Age, menarche, parity, age at first birth, BMI (current and age 18), breastfeeding, oral contraceptive use, menopausal status, breast biopsy, first-degree family history of BC | Age, menarche, age at first birth, parity, height, BMI, alcohol, smoking, oral contraceptive use, HRT, age at menopause, benign breast disease, first-degree family history of BC |
| Model risk factors excluded from current analyses | Atypical hyperplasia | BRCA1/2, family history (2nd degree), PRS; Atypical hyperplasia, mammographic density | Full pedigree family history, BRCA1/2/PALB2/CHEK2/ATM pathogenic variants, 313-SNP PRS Mammographic density | Family history of prostate cancer; bilateral oophorectomy | PRS; Mammographic density; |
| Missing data | Hard rules | Excluded | Marginalized via pedigree likelihood integration | Hard rules and excluded | Imputed using a reference population |

To evaluate the role of incidence-rate assignment in model performance, we compared AUCs and E/O ratios for the iCARE-Lit model using SEER incidence rates for non-Hispanic White individuals vs. Black individuals in two cohorts with extremely different racial makeups: BWHS (100% Black women) and the Agricultural Health Study (AHS, 96% White women).

## RESULTS

### Descriptive Characteristics

A total of 1,591,803 women ages 20-75 years were considered in this analysis, representing 21 cohorts in the United States, Canada, the United Kingdom, Norway, and Australia (**Table 2**).

**Table 2.** Characteristics of 21 Cohorts in the Breast Cancer Risk Prediction Project (BCRPP) included in analysis^1^.

| Cohort<sup>2</sup> | Acronym | Country | Total N (%) | Prevalent case or case 0-<1y<sup>3</sup><br>N (%) | Invasive BC 1-5y<sup>4</sup>, N (%) | Invasive BC 2-6y<sup>5</sup>, N (%) | Study enrollment | Birth year, median (range) |
| --- | --- | --- | --- | --- | --- | --- | --- | --- |
| **All Cohorts** |  |  | **1,591,803** | **3,110** | **20,760** | **21,627** | - | - |
| The Agricultural Health Study | AHS | U.S. | 33,393 (2.1%) | 28 (0.9%) | 375 (1.8%) | 384 (1.8%) | 1993-1997 | 1949 (1918-1977) |
| Breast Cancer Now Generations Study | BGS | U.K. | 102,567 (6.4%) | 118 (3.8%) | 1,158 (5.6%) | 1,190 (5.5%) | 2004-2011 | 1930 (1920-1940) |
| Black Women's Health Study | BWHS | U.S. | 44,685 (2.8%) | 0 (0.0%) | 512 (2.5%) | 510 (2.4%) | 1995 | 1956 (1927-1974) |
| The Beta-Carotene and Retinol Efficacy Trial | CARET | U.S. | 6,289 (0.4%) | 20 (0.6%) | 138 (0.7%) | 145 (0.7%) | 1985 | 1933 (1917-1944) |
| Cancer Prevention Study 2 | CPS2 | U.S. | 97,222 (6.1%) | 71 (2.3%) | 1,738 (8.4%) | 1,798 (8.3%) | 1982 | 1930 (1916-1952) |
| Cancer Prevention Study 3 | CPS3 | U.S. | 232,802 (14.6%) | 344 (11.1%) | 2,513 (12.1%) | 2,620 (12.1%) | 2006-2013 | 1963 (1933-1991) |
| California Teacher's Study | CTS | U.S. | 114,780 (7.2%) | 99 (3.2%) | 1,884 (9.1%) | 2,050 (9.5%) | 1995-1996 | 1940 (1920-1980) |
| Canadian Study of Diet, Lifestyle, and Health<sup>6</sup> | CSDLH | Canada | 3,503 (0.2%) | 22 (0.7%) | 356 (1.7%) | 368 (1.7%) | 1992-1998 | 1941 (1918-1975) |
| JANUS | JANUS | Norway | 145,927 (9.2%) | 627 (20.2%) | 804 (3.9%) | 902 (4.2%) | 1974-2016 | 1947 (1912-1972) |
| Melbourne Collaborative Cohort Study | MCCS | Australia | 13,976 (0.9%) | 713 (22.9%) | 208 (1.0%) | 198 (0.9%) | 2003-2007 | 1940 (1920-1960) |
| The Multiethnic Community Cohort | MEC | U.S. | 105,601 (6.6%) | 164 (5.3%) | 1,815 (8.7%) | 1,874 (8.7%) | 1993-1996 | 1934 (1918-1953) |
| Mayo Mammography Health Study<sup>6</sup> | MMHS | U.S. | 3,214 (0.2%) | 53 (1.7%) | 229 (1.1%) | 256 (1.2%) | 2003-2006 | 1948 (1927-1971) |
| Nurses' Health Study | NHS | U.S. | 121,529 (7.6%) | 78 (2.5%) | 826 (4.0%) | 855 (4.0%) | 1976 | 1933 (1921-1946) |
| Nurses' Health Study 2 | NHS2 | U.S. | 116,412 (7.3%) | 34 (1.1%) | 383 (1.8%) | 424 (2.0%) | 1989 | 1954 (1945-1964) |
| New York University Women's Health Study | NYUWHS | U.S. | 14,273 (0.9%) | 70 (2.3%) | 235 (1.1%) | 251 (1.2%) | 1985-1991 | 1935 (1915-1956) |
| Prostate, Lung, Colorectal, and Ovarian Cancer Screening Trial | PLCO | U.S. | 76,110 (4.8%) | 74 (2.4%) | 1,510 (7.3%) | 1,550 (7.2%) | 1993-2001 | 1934 (1919-1948) |
| Southern Community Cohort Study | SCCS | U.S. | 48,984 (3.1%) | 49 (1.6%) | 554 (2.7%) | 558 (2.6%) | 2002-2009 | 1954 (1927-1969) |
| Sister Study | SIS | U.S. | 50,794 (3.2%) | 173 (5.6%) | 1,323 (6.4%) | 1,339 (6.2%) | 2004-2009 | 1951 (1928-1974) |
| U.S. Radiologic Technologists Cohort | USRT | U.S. | 70,790 (4.4%) | 37 (1.2%) | 414 (2.0%) | 458 (2.1%) | 1983-1989 | 1949 (1908-1966) |
| Women's Health Initiative | WHI | U.S. | 149,295 (9.4%) | 271 (8.7%) | 3,187 (15.4%) | 3,253 (15.0%) | 1993-1998 | 1931 (1919-1945) |
| Women's Health Study | WHS | U.S. | 39,657 (2.5%) | 65 (2.1%) | 598 (2.9%) | 644 (3.0%) | 1993-1995 | 1941 (1917-1955) |

1 Restricted to individuals ≥20 and <75 y of age at baseline record
2 Listed alphabetically by cohort acronym
3 Considered prevalent case at baseline if year of dx (invasive or *in situ* BC) ≤ year of the first record
4 Excludes prevalent cases; considered a case if diagnosis year ≥ first record year + 1 and non-missing year of diagnosis
5 Excludes cases within 1-y of record date; considered a case if diagnosis year ≥ first record year +2 and non-missing year of diagnosis
6 Harmonized data provided to BCRPP as case-cohort selection. Cases diagnosed after 1-5y or 2-6y that were not part of the original subcohort were excluded from respective analyses.

After excluding 3,110 prevalent cases, 1,588,693 individuals remained, among whom, 20,760 (1.3%) were diagnosed with invasive BC within the first five years of follow-up (**Table 2**).

Descriptive characteristics across cohorts are provided in **Table S2**.

Several cohorts were missing risk factor variables relevant to one or more of the models tested, due to either lack of collection on baseline surveys or lack of harmonization for BCRPP (**Figure 1**). Variables with high missingness by design included history of BBD, biopsy number, and detailed family history information. All cohorts provided information on height, BMI, menopausal status, and first-degree family history (yes/no) of BC.

### Absolute risk distribution

Distribution of absolute 5-year invasive BC risk varied between cohorts, with median risks ranging from 0.28% (NHS2, BOADICEA model) to 3.29% (SIS, iCARE-Lit model) (**Table S3**). Within individual cohorts, absolute risk distributions were similar across different models (**Table S3, Figure 2**). Lower risks were seen among cohorts with younger baseline ages. A narrower range of absolute risk was observed in cohorts with a greater proportion of missing risk factor data (e.g., CARET, MCCS), and with limited age distributions (**Figure 2**). The range of absolute risk estimates was wider for iCARE-Lit and TC models.

**Figure 2.**
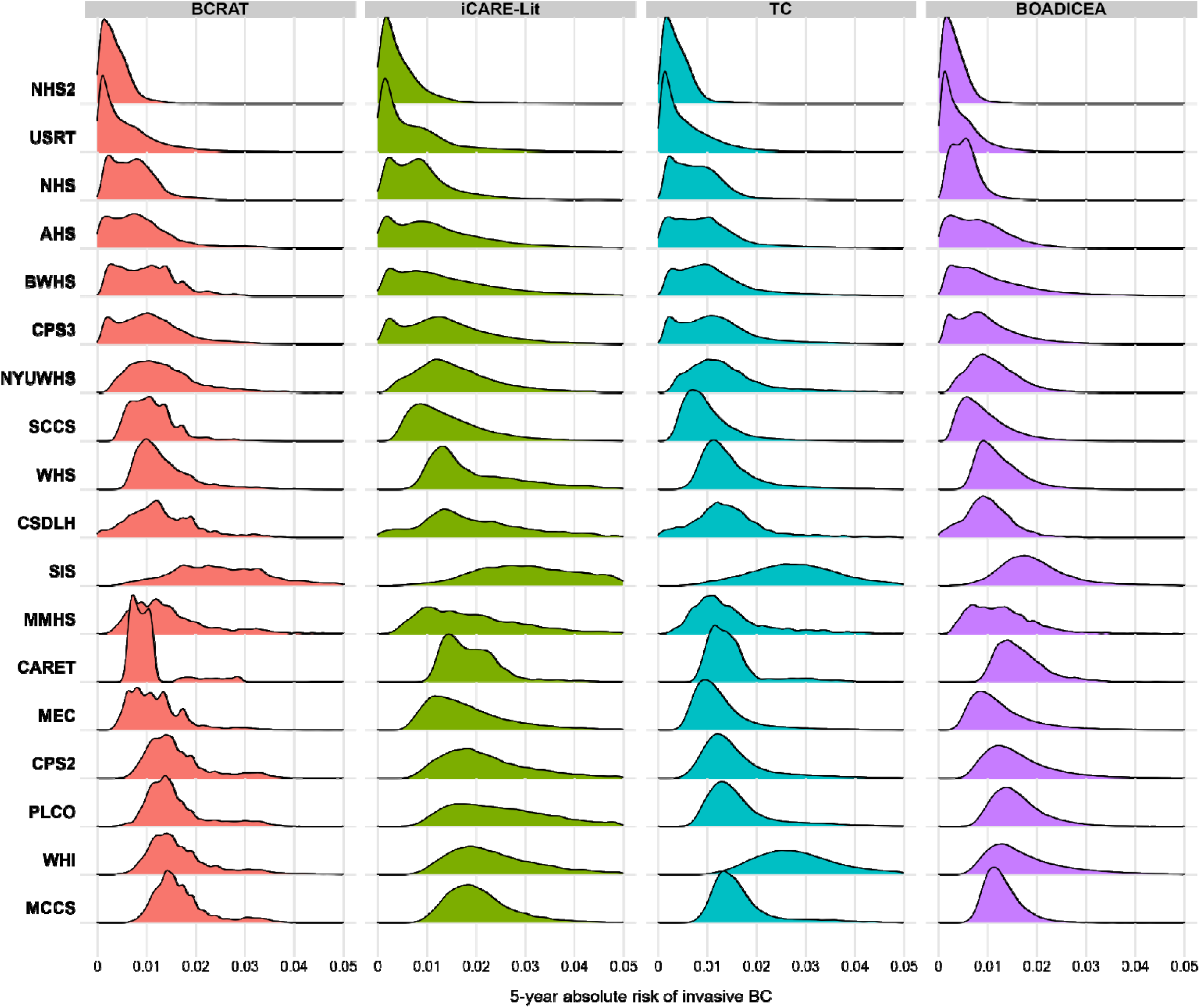
Distribution of predicted 5-year absolute risk by model and cohort among cohorts with individual-level data. Cases are those that developed invasive BC 1-5 years from baseline questionnaire. For visualization purposes, cohorts are ordered by mean age from youngest cohort (NHS2, mean age=34.4 y) to oldest cohort (WHI, mean age=63.4 y) and maximum risk set at 0.05 to capture 0-90^th^ percentile risk for all cohorts (e.g., SIS, 90^th^ %tile of risk=0.044). Minimum, Q1, Q2, Q3, and maximum absolute risks are given for all cohorts in Table S3. BCRAT: Breast Cancer Risk Assessment Tool; iCARE-Lit: Individualized Coherent Risk Estimation (iCARE) Literature Model; TC: Tyrer-Cuzick model; BOADICEA: The Breast and Ovarian Analysis of Disease Incidence and Carrier Estimation Algorithm. Genetic data was not available for models.

### Model Discrimination

After age-adjustment, discrimination was consistent between cohorts (**Figure 3**, **Table S4**). Before age adjustment, higher AUCs in younger cohorts (**Figure S1**) reflect the known strong relationship between age and BC risk. Average discrimination was lowest for BCRAT (pooled AUC [95% CI]=0.57 [0.56-0.57]), while other models had equivalent average AUCs of 0.58 (0.57-0.59) (**Figure 3**). Between-study heterogeneity was significant in all models (p<0.001), though proportion of variance explained by cohort heterogeneity varied by model [I^2^ = 61.7% (BCRAT), 73.1% (iCARE-Lit), 69.3% (TC), 64.9% (BOADICEA)]. Associations between cohort characteristics and discrimination were minimal and inconsistent across models (**Table S5**).

**Figure 3.**
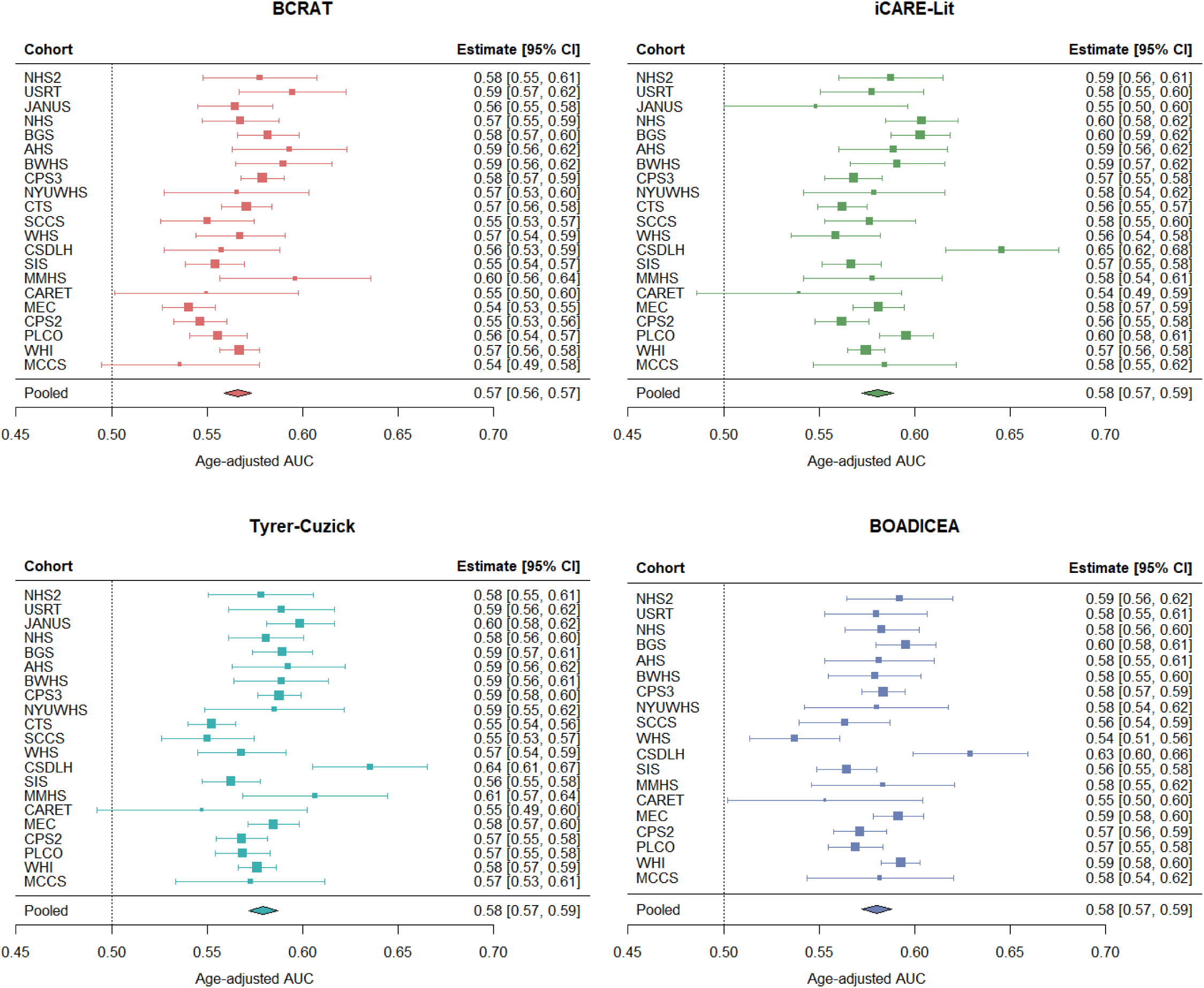
Age-adjusted AUC based on absolute risk by model for all cohorts. Pooled estimates are given from random effects meta-analysis. AUC was calculated within cohort-specific age quintiles and combined AUC weighted by the inverse of the variance for each quintile. Cohorts appear in order of mean age at baseline questionnaire (youngest to oldest from top to bottom). BCRAT: Breast Cancer Risk Assessment Tool; BOADICEA: The Breast and Ovarian Analysis of Disease Incidence and Carrier Estimation Algorithm; iCARE-Lit: Individualized Coherent Risk Estimation (iCARE) Literature Model. Note BOADICEA was not calculated for JANUS and CTS. I^2^ (total heterogeneity/total variability) BCRAT: 60.3%, iCARE-Lit: 73.1%, Tyrer-Cuzick: 69.3%, BOADICEA: 64.9%. P-heterogeneity <0.001 for all models. Note, the estimate for iCARE-Lit for JANUS included only postmenopausal women (N=12,333) as there were a limited participants with complete information on covariates among premenopausal women, precluding creation of a premenopausal reference dataset for this cohort.

### Model Calibration

Across all models and cohorts, calibration was less consistent than AUC (**Table S6, Figure 4, Figure S2**), with average E/O ratios ranging from 0.47 to 3.03. Meta-analyses demonstrated significant underestimation by BOADICEA (pooled E/O=0.81 [0.76-0.86]) and overestimation by iCARE-Lit (pooled E/O=1.25 [1.13-1.37]) (**Figure S2**). The test for between-study heterogeneity was significant in all models (p<0.001, I^2^>97% for all). While average E/O ratios were near one (|E/O-1| ≤0.30) in 70% of model-cohort combinations (**Table S7**), deviation was pronounced at the tails of the distribution of 5year risk. Notably, among individuals who were categorized as high-risk (≥3% 5-year risk), overestimation was common for all models (**Figure 4**). The highest overestimation across cohorts with N>250 participants in the high-risk category was in iCARE-Lit (pooled E/O=1.60 [1.45-1.76]).

**Figure 4.**
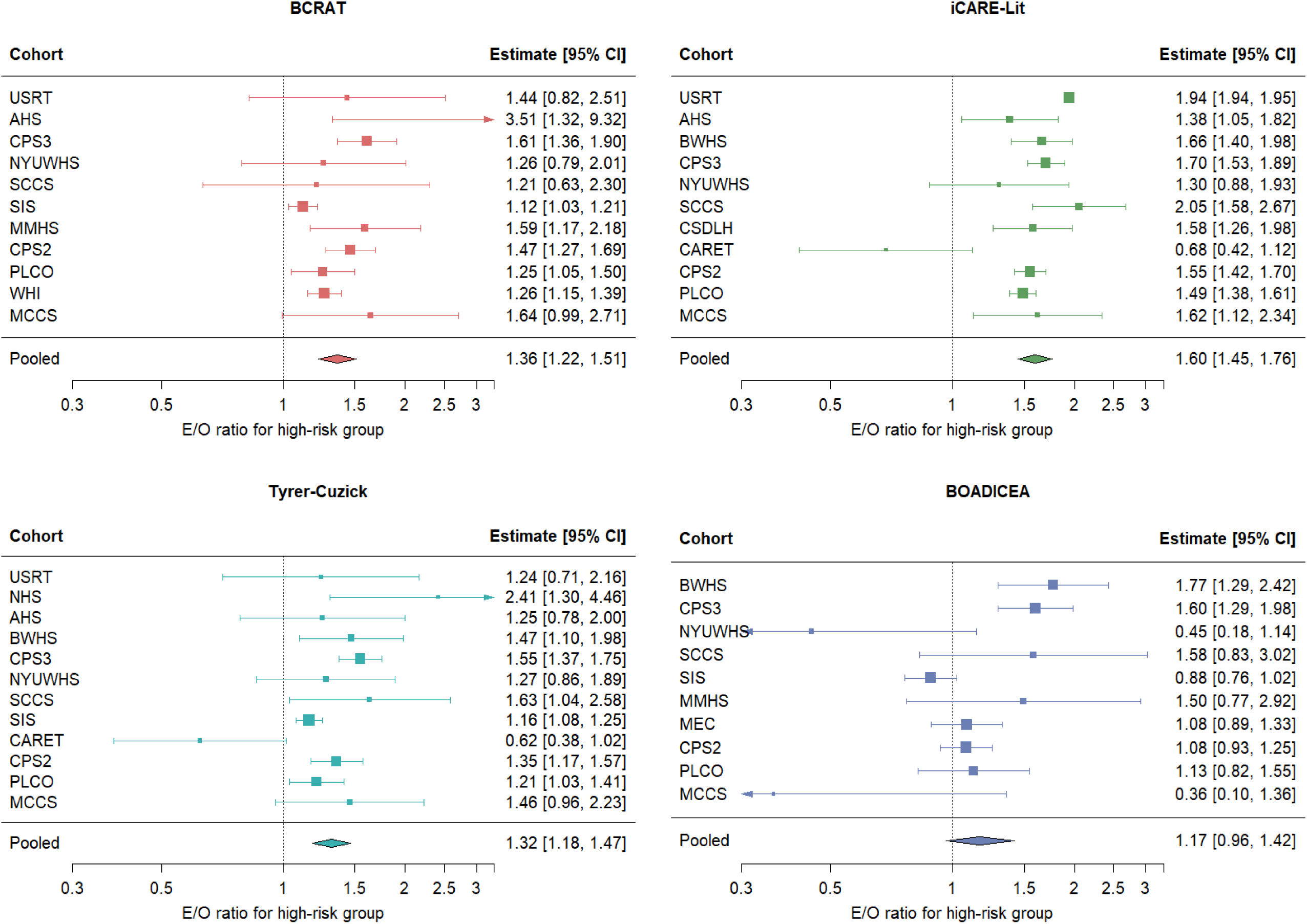
Expected to observed (E/O) ratios for participants considered high-risk (absolute 5-year risk ≥3.0%). Cohorts are listed by age at baseline from lowest to highest. E/O ratios are shown on the log scale. Results not shown for cohorts with effective sample size <250. Results are not given for BGS, CTS, and JANUS due to data sharing limitations.

**Figure 5.**
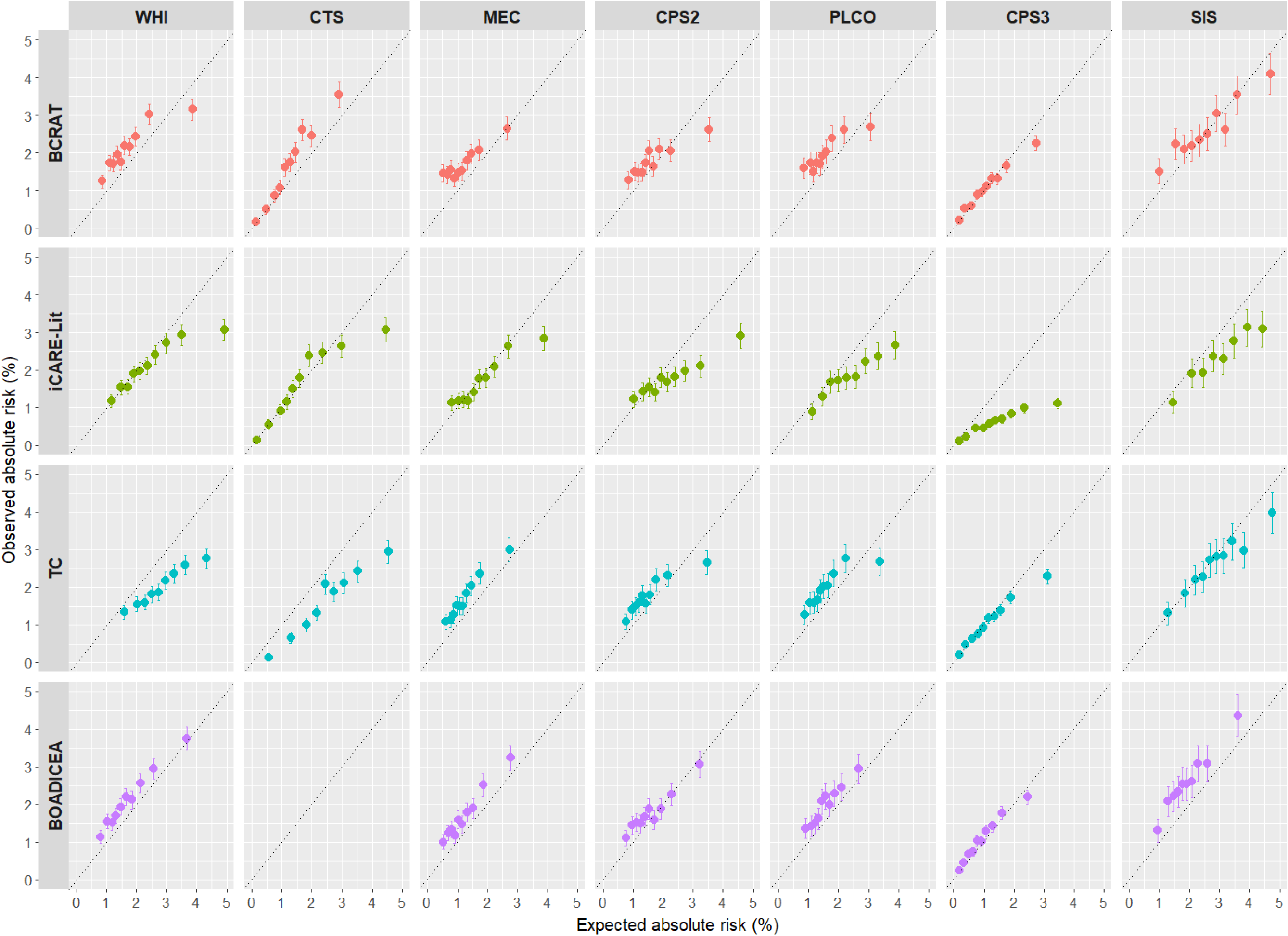
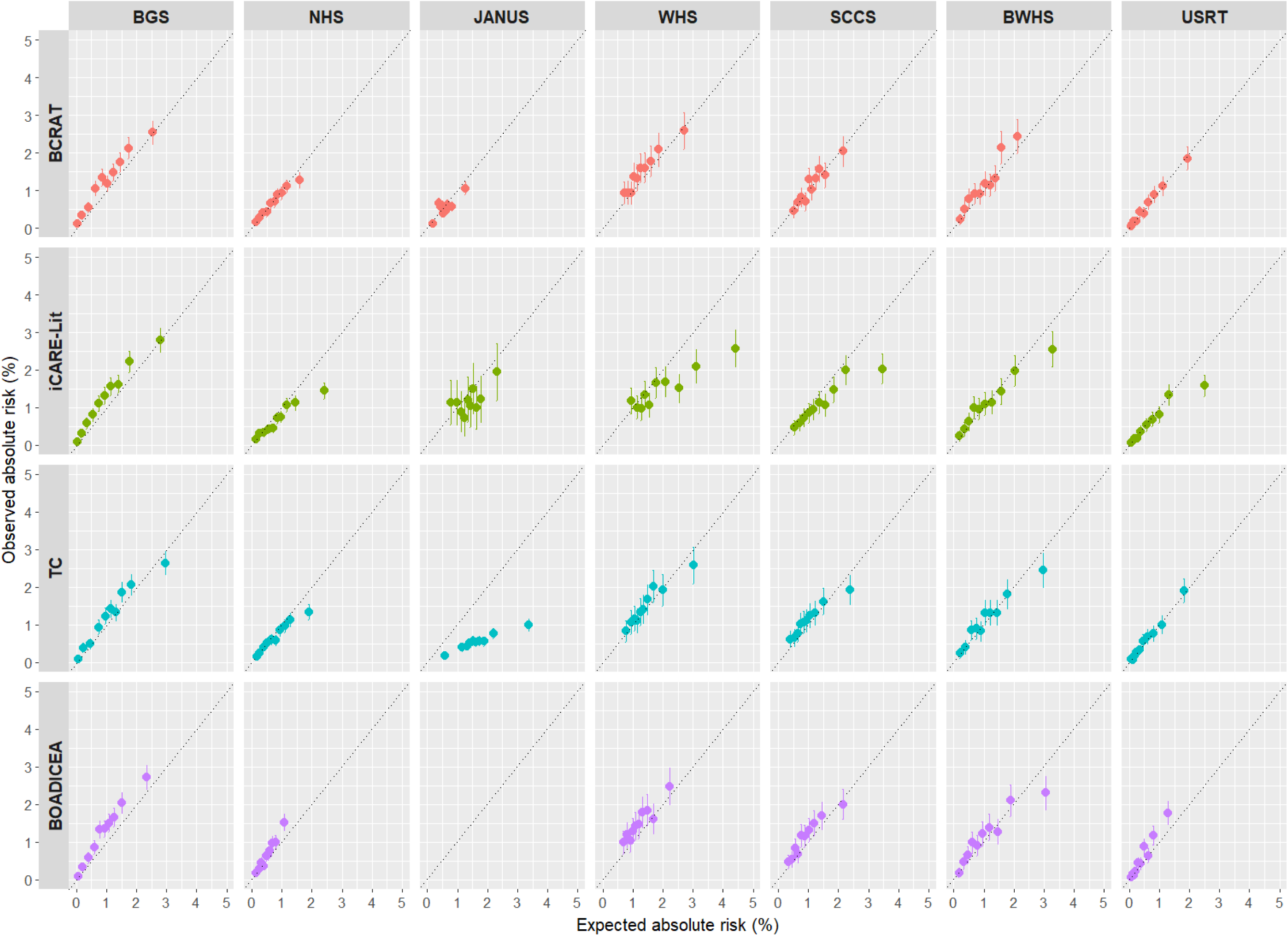
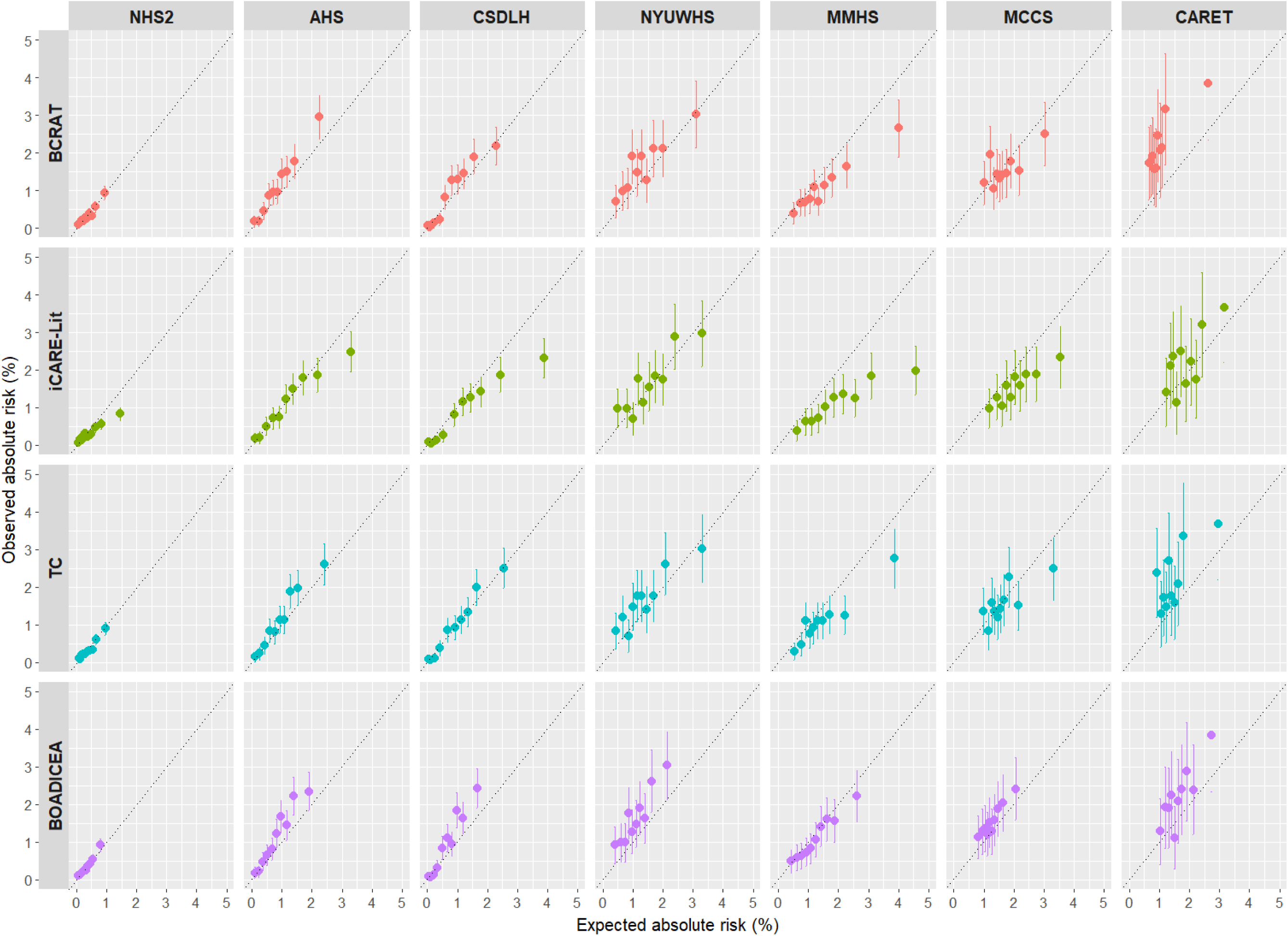
Observed (95% CI) vs. expected absolute risk by model and cohort, by decile of cohort-specific predicted risk. Cohorts are ordered from high to low by number of breast cancer cases in 5 years. BCRAT: Breast Cancer Risk Assessment Tool; BOADICEA: The Breast and Ovarian Analysis of Disease Incidence and Carrier Estimation Algorithm; iCARE-Lit: Individualized Coherent Risk Estimation (iCARE) Literature Model; TC: Tyrer-Cuzick model. Graphs are truncated at 5% to allow for visual comparison across deciles (the 10^th^ decile of SIS for iCARE-Lit model is not pictured, see Table S6). Note BOADICEA was not calculated for JANUS and CTS.

Associations of cohort characteristics with E/O ratios were, like AUC associations, small and inconsistent. Deviations in E/O ratios were associated with later cohort birth year (iCARE-Lit, slope=0.038, p=0.004), and less information on number of first-degree relatives with BC (BCRAT, slope=-0.009, p=0.009).

Calibration patterns were more similar across cohorts and models when restricted to cohorts that had all relevant model variables (**Figure S3**).

### Model performance among Black Women

The performance of the BWHS RC was tested in four studies, restricted to Black women: BWHS (N=43,185, N=493 cases in 5y), the Women’s Health Initiative (WHI) (N=12,089, N=195 cases in 5y), the multiethnic cohort (MEC) (N=17,333, N=277 cases in 5 y), and the Southern Community Cohort Study (SCCS) (N=30,492, N=343 cases in in 5 y). Absolute risks were similar between cohorts (median [IQR], BWHS: 1.0% [0.6-1.4%] vs. WHI: 1.2% [1.0%-1.5%] vs. MEC: 1.3% [2.2%-2.7%] vs. SCCS: 1.0 [0.7-1.3%], p-value for Kruskal-Wallis test=0.39) (**Table S8**). Prior to additional age-adjustment, discrimination based on absolute risk was higher in BWHS (AUC [95% CI]=0.66 [0.64-0.69]) compared to other cohorts (AUC range=0.57-0.61) (**Table S9**). After age adjustment, model performance was similar across cohorts (AUC range=0.55-0.59) (**Table S9, Figure S4**). Discrimination and calibration were consistent across risk models within cohorts (**Figure 6, Tables S9-11**).

**Figure 6.**
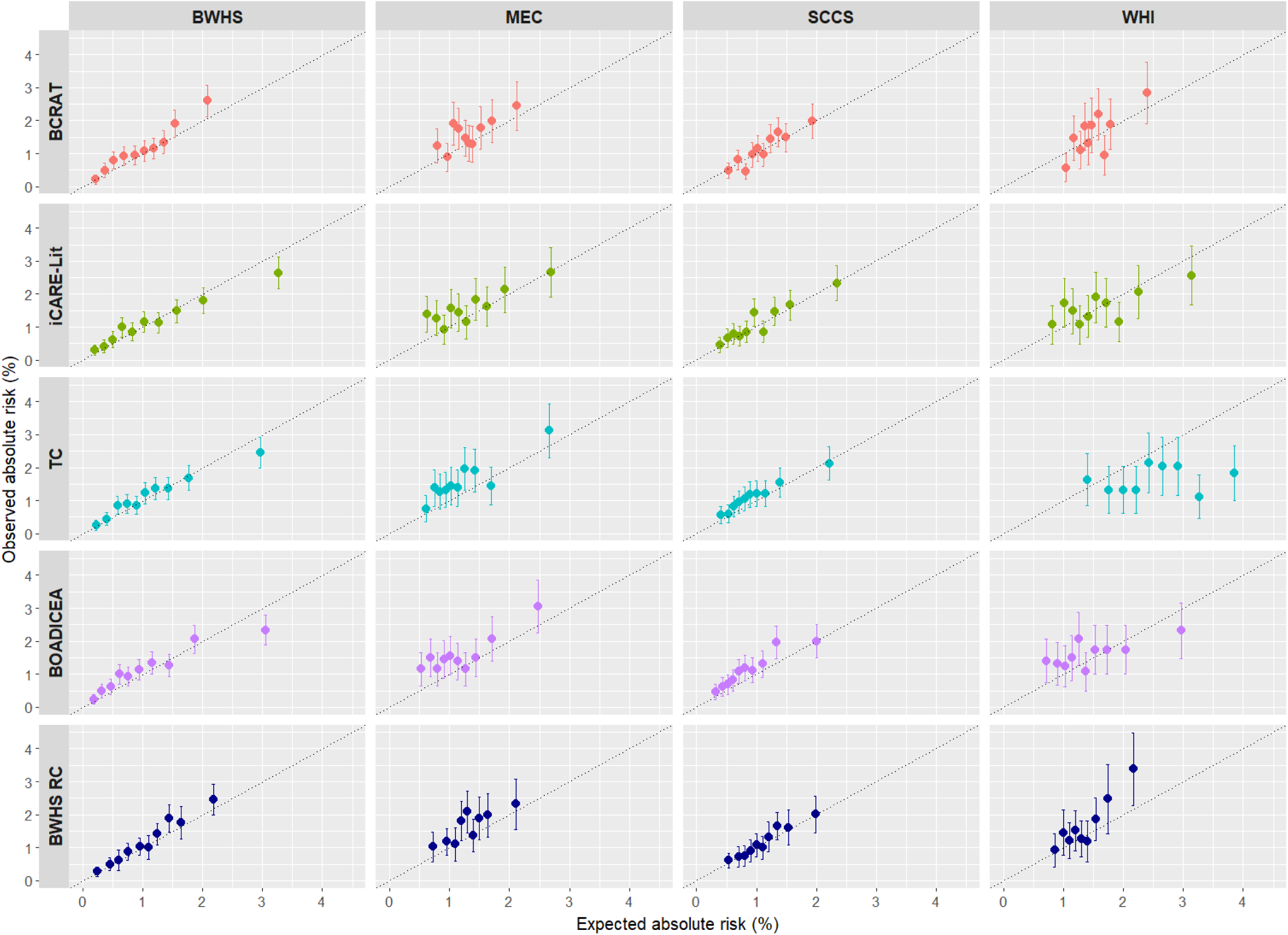
Observed to expected 5-y absolute risk of invasive BC among Black women aged 30-70 years in BCRPP comparing risk models. Note: the BWHS RC model includes prostate cancer history, which was not collected for data harmonization and treated as missing (set to the lowest risk category) for all cohorts. Cohorts compared include those with >100 invasive BC cases in Black women in 5-years. BCRAT: Breast Cancer Risk Assessment Tool; BOADICEA: The Breast and Ovarian Analysis of Disease Incidence and Carrier Estimation Algorithm; iCARE-Lit: Individualized Coherent Risk Estimation (iCARE) Literature Model; TC: Tyrer-Cuzick model. BWHS RC: Black Women’s Health Study Breast Cancer Risk Calculator; BWHS: Black Women’s Health Study; WHI: Women’s Health Initiative; SCCS: Southern Community Cohort Study; MEC: Multiethnic Community Cohor

### Incidence rate assignments

Applying incidence rates from more representative populations improved calibration of the iCARE-Lit model but did not influence discrimination. For BWHS (100% Black women), if using rates from non-Hispanic Black women, average E/O was 1.02 (0.94-1.12), compared to 1.32 (1.21-1.44) when using rates for non-Hispanic White women. For AHS (96% White women), E/O was 1.08 (0.98-1.20) applying rates from non-Hispanic White women but 0.84 (0.76-0.93) when using rates from Black women.

All results were virtually unchanged when assessing follow-up from year two.

## DISCUSSION

In this comprehensive comparison of BC risk models using classical risk factors in 21 cohorts with BCRPP-harmonized data, we found similar age-adjusted discrimination across cohorts and a tendency to overestimate risk of women with >3% 5-year risk. No clear trends were observed for this set of cohorts in performance metrics by baseline age, birth year, and racial diversity.

Poor calibration at the tails of risk deciles suggests potential departures from the multiplicative model. As the largest study to date to systematically evaluate and compare model performance of existing BC risk prediction models in women across the U.S., Canada, Europe, and Australia, including appreciable numbers of U.S. Black women, evidence from this study supports the development of a unified risk model to provide robust risk prediction across diverse populations, rather than population-specific models.

Average model discrimination estimates (median age-adjusted AUCs: 0.56-0.58) align well with previous model validation efforts in several cohorts for BCRAT,^7,19^ iCARE-Lit,^13^ TC,^16^ BOADICEA,^20^ and the BWHS RC.^14,21^ A systematic review and meta-analysis of cohorts noted major differences in discriminatory ability and model calibration based on population, with lower performance observed in non-European cohorts.^22^ In the pooled BCRPP analyses, we found age-adjusted model discrimination to be similar across cohorts, indicating that covariate distributions conditional on age were likely similar. With respect to calibration, poor performance at the tails of risk distributions, with consistent overestimation of risk for high-risk individuals seen across most BCRPP cohorts, was previously observed when comparing iCARE-Lit performance across 15 different cohorts (N=8 in BCRPP, N=7 unique).^13^

Previous studies have sought to directly compare the performance of specific models, though none were as large or as diverse as BCRPP. The performance of BCRAT and TC were compared in a mammography cohort (2007-2009) of 35,921 women aged 40-84, where average model calibration was better using BCRAT (O/E=0.98) vs. TC (O/E=0.84).^23^ We also observed greater overestimation of risk using the TC (vs. BCRAT) model here, though this was not universal. Others have shown that BCRAT underestimates risk among individuals with a family history of BC, while TC overestimates risk.^24^ However, here we showed that, within a cohort with 100% positive family history of BC (Sister Study), performance was similar for BCRAT and TC.

Population-average age-specific cancer incidence rate assignment had a large impact on model calibration and was more influential than relative risk assignments, demonstrating the need to use incidence rates representative of the target population. For example, after assigning race-specific incidence rates, calibration metrics were similar for the iCARE-Lit model vs. the BWHS RC among Black women, despite the development of the BWHS RC entirely in Black women and derivation of relative risk estimates for iCARE-Lit from majority White women. This, along with our finding that cohort racial diversity measured by GSDI did not influence model performance, suggests that age-specific relative risk for individual risk factors may not differ substantially by race, which may support the use of a unified model across diverse populations with proper incidence rate assignment. However, our primary comparisons are limited to White vs. Black women and GSDI does not capture all intricacies relevant to racial/ethnic distribution. Further support for a unified model is indicated by our finding of no consistent trends between cohort characteristics, such as age and birth year, and model performance.

The variability in the proportion of missingness for key risk factors across cohorts may account for some of the variability in model performance metrics seen here, as higher variable missingness tended to result in lowered AUCs in meta-regressions. However, most associations were not significant; while missingness of variables with higher relative risks (e.g., biopsy and BBD history) shifted some metrics, this was inconsistent across models. Despite suspected minimal impact of missingness on model performance, our results are not necessarily generalizable to the ideal situation in which individuals have information on all risk factors.

Our study has several limitations. Because we did not collect two risk factors used in the BWHS BC Risk Calculator, family history of prostate cancer and bilateral oophorectomy, risk may be underestimated in some individuals. When applying the iCARE-Lit model, we used the same reference populations for all U.S. cohorts which may misrepresent risk factor distributions of some cohorts. Our assumption that first-degree family history of BC meant the mother had BC (if not specifically reported) may have led to overestimations in expected risk for the TC and BOADICEA models; however, we found estimated risk differences to be minimal when testing alternative assumptions. While our primary comparison included women aged 20-75 years at baseline entry to capture the largest population across models, the BCRAT model is not recommended for use in individuals <35 years of age. Some misclassification is likely in younger individuals, though it is likely to be similarly misclassified in all models, as none were developed or validated in populations with large numbers of young women. In this analysis, 17,403 women (1.4%) were aged 20-35 years, accounting for only 0.2% of 5-year BC cases. It is unlikely the inclusion of these individuals substantially altered model performance metrics; however, our results are not generalizable to the youngest women. While we sought racial diversity for this project, only four cohorts had a sufficient number of cases (195 to 493) to assess performance in Black women and representation of individuals from Africa, Asia, and Central and South America was lacking. In addition, representation from Europe was low relative to the U.S., further limiting generalizability. Finally, while we were able to assess the stability of model performance over the range of birth cohorts captured in these studies, recent changes in exposures and risk factor distribution in early and middle life (e.g. changes in age at menarche, age at first birth, GLP-1 usage) may affect model performance and will need to be evaluated in newer cohorts.

Overall, our findings highlight consistencies of model discrimination despite differences in cohort demographics, as well as overestimation of risk for high-risk individuals when using classical risk factor data. This overestimation has clinical implications as these individuals may receive unnecessary chemoprevention. Our finding that model performance was not altered substantially by cohort demographics suggests that a unified model may sufficiently capture risk across diverse populations and emphasizes the importance of representative absolute incidence rate assignment. Through BCRPP, we propose development and evaluation of a new, comprehensive model that enables estimation in diverse populations and incorporates more complex risk factor relationships to improve calibration at the high and low ends of risk. Given the low discrimination of models using classical risk factors only, the new model will also require mammographic density and genetic risk, which have been shown to enhance discrimination.^25,26,26^ As we work to develop better models that can be used in prevention settings as well as in clinic, it is essential to consider feasibility, teachability of tools and ease of interpretation, and risk-benefit analyses.^27–29^

## Supporting information

Funding information and author contributions

## Data Availability

All data produced in the present work are contained in the manuscript and its supplemental materials. Data access for BCRPP is facilitated through the BCRPP Data Platform in accordance with data transfer agreements signed with participating studies and the policies and procedures of the BCRPP Data Coordinating Center (DCC) at the Division of Cancer Epidemiology and Genetics at the National Cancer Institute.

## Ethics approval

All participating cohorts included were approved by their respective IRBs. This study conforms to the standards of the Declaration of Helsinki.

## Author contributions

Author contributions are available as a Supplementary file online.

## Supplementary Data

Supplementary data are available online.

## Data Availability

Data access is facilitated through the BCRPP Data Platform (https://epidataplatforms.cancer.gov/bcrpp/) in accordance with data transfer agreements signed with participating studies and the policies and procedures of the BCRPP Data Coordinating Center (DCC) at the Division of Cancer Epidemiology and Genetics at the National Cancer Institute. Researchers who are interested in using BCRPP data can submit a study concept describing the project through the Data Platform. The data access coordinating committee (DACC) will review, and if a concept is approved, individual studies participating in BCRPP are notified and given a time period to opt-out from the approved project. Before work can begin, a data transfer agreement must be signed between the researcher’s institution and the BCRPP DCC. Access is only granted for purposes outlined in approved proposals. Data cannot be downloaded or saved on local computers or any persistent storage device other than the controlled-access workspace provided by the BCRPP. Note that data from the European JANUS cohort are protected by the principles set out in articles 6 (1) I and 9 (2) (j) of the General Data Protection Regulation (GDPR), and Norwegian law. Requests to access these data can be directed to the co-authors representing the JANUS cohort and can be facilitated after obtaining the necessary ethical and legal approvals. To request the ability to submit study concepts to the BCRPP DACC, please. Note that study concepts submitted to the BCRPP DACC should not request access to data from only a few cohorts that are participating in the BCRPP.

## Use of Artificial Intelligence

Artificial intelligence was not used at any point when preparing this manuscript

