## Supplemental Methods for "Performance of general-population breast cancer risk prediction models in an international consortium"

**Description of Cohorts participating in BCRPP**

There are currently 26 cohorts that have agreed to contribute to BCRPP in model development efforts, representing the U.S. (N=18), Europe (N=6), Australia (N=1), and Canada (N=1). In addition, 2 U.S.-based cohorts have also agreed to support model validation efforts, the Breast Cancer Surveillance Consortium (BCSC)^1^ and Vitamin D and Omega 3 Trial (VITAL)^2^. For this analysis, 21 cohorts participated, from the U.S. (N=17), Europe (N=2), Australia (N=1) and Canada (N=1). Below we have summarized cohort characteristics for the 21 BCRPP studies included in our analysis.

Agricultural Health Study (AHS)

The AHS was started in 1993 and funded by the National Cancer Institute and the National Institute of Environmental Health Sciences to answer questions about the health of farming populations. The study enrolled 52,394 private pesticide applicators and 32,345 of their spouses from Iowa and North Carolina between 1993-1997. At baseline, participants answered detailed questionnaires collecting information on demographics, health status, pesticide exposure, and more.^3^ To date, there have been four rounds of follow-up (1999-2003, 2005-2010, 2013-2015, 2019-2021), during which participants were asked to update information on pesticide exposure and medical history. In the first round of follow-up (1999-2003), participants also answered diet questionnaires and provided buccal cells for research purposes. Over the course of the study there are regular linkages to cancer registries, mortality registries, and Medicare claims data. Details on the study are found online: https://aghealth.nih.gov/about/.

Breast Cancer Now Generations Study (BGS)

The BGS is a prospective cohort started in 2003 with the goal of better understanding breast cancer development, prognosis, and life after breast cancer. From 2004-2011 over 110,000 women were enrolled. At enrollment, individuals completed a recruitment questionnaire that included information participant demographics, lifestyle, reproductive history, anthropometrics, medical history, family history of cancer, and other factors known or suspected to be linked to cancer risk. Blood samples were collected at recruitment for 92% of participants for biomarkers studies. Participants are asked to update their exposures and health history on follow-up questionnaires, which have been sent 2, 6, 9, and 13 years after recruitment to date. Cancer diagnoses are self-reported and linked to national cancer registries. Mortality information is collected by family report and linkage to death registries.^4^ Additional details are found online: https://breastcancernow.org/our-research/research-centres-and-projects/individual-research-projects/the-generations-study.

Black Women’s Health Study (BWHS)

The BWHS was established with the goal to improve health among U.S. Black women by understanding risk factors for and management of diseases relevant to Black women. In 1995, 59,000 self-identified Black women aged 21-69 years were enrolled, at which time they completed a baseline questionnaire providing reproductive, lifestyle, anthropometric, diet, and other factors. Biennial follow-up questionnaires collect data on disease status and changes in risk factor exposures from these women. Saliva, blood, and tumor tissue samples were collected for a subset of the cohort. Incident breast cancer diagnoses are identified by self-report and confirmed by medical record review or linkage to cancer registries and the National Death Index.^3^ Additional details are found online: https://www.bu.edu/bwhs/.

Carotene and Retinol Efficacy Trial (CARET)

CARET was a randomized double-blind trial to test the safety and efficacy of using beta-carotene and retinyl palmitate in individuals at high risk of lung cancer. Recruitment began in 1985, and participants were recruited from six study centers in Seattle, WA (coordinating center), Baltimore, MD, Irvine, CA, Groton, CT, Portland, OR; San Francisco, CA. The intervention was stopped in 1996 after the study found a harmful effect though active follow up continued through 2005, with passive follow-up conducted through linkages to the National Death Index and cancer registries through 2013. At baseline and during the study, the study collected data on smoking history, medical conditions, family cancer history, dietary intake, occupational history of asbestos exposure, and demographics. During the intervention, individuals also provided serum, plasma, whole blood, and lung tissue specimens that make up the Biorepository.^6,7^ Additional details and publications are found online: https://www.fredhutch.org/en/research/divisions/public-health-sciences-division/research/cancer-prevention/carotene-and-retinol-efficacy-trial.html.

California Teachers Study (CTS)

The CTS recruited female teachers and administrators who were active members of the California State Teachers' Retirement System (CalSTRS) in 1995, enrolling 133,476 women between 1995-1996. Participants completed baseline questionnaires with detailed information on demographics, health, and lifestyle factors. Full length follow-up surveys were completed in 1999, 2001, 2006, 2013, and 2018.^8^ Biospecimens, including blood, saliva, urine, and buccal cells, have been collected at various follow up points on subset of CTS participants. Linkages to the California Cancer Registry provide details on diagnosis, including type of cancer, stage, grade, and subtype. Mortality information is captured via linkage to national and state mortality data and include information on date and cause of death. CTS additionally collects information on inpatient hospitalizations via linkage to State data. See the website for more details: https://www.calteachersstudy.org/.

Cancer Prevention Study 2 (CPS2)

CPS2 is a prospective cohort study led by the American Cancer Society (ACS) started in 1982 and included 1.2 million American men and women recruited by ACS volunteers. Participants completed a baseline questionnaire including personal and family history of diseases, demographics, anthropometrics, reproductive history, diet, and lifestyle factors. Continued follow-up of participants has been conducted via the CPS2 Nutrition Surveys, which were developed to obtain more detailed diet information and to allow for prospective cancer incidence follow-up. Questionnaires collecting risk factor exposures, diet, and cancer outcomes, were sent to 184,000 women in 1992 and every two years from 1997-2015. Self-reported cancers are confirmed by medical record and linkage with state cancer registries. Biological specimens, including blood and cheek cell samples, have also been collected from a subset of the cohort. The full cohort is followed for mortality outcomes. Cohort information can be found on the ACS website: https://www.cancer.org/research/population-science/cancer-prevention-and-survivorship-research-team/acs-cancer-prevention-studies.html

Cancer Prevention Study 3 (CPS3)

Over 300,000 men and women in the US and Puerto Rico between ages 30-65 years were enrolled in CPS3 between 2006-2013. Approximately 17% of participants are racial minorities. At baseline, participants completed questionnaires containing detailed lifestyle, demographic, and environmental data, and 99% of participants provided blood samples. The first full follow-up survey with detailed diet questions and tumor tissue collection was completed in 2015, with others completed in 2021, 2024, and every three years thereafter. Cohort information can be found on the ACS website: https://www.cancer.org/research/population-science/cancer-prevention-and-survivorship-research-team/acs-cancer-prevention-studies.html

Canadian Study of Diet, Lifestyle, and Health (CSDLH)

The CSDLH was established to study the relationship between lifestyle factors, biomarkers, and cancer incidence. Between 1992-1998 73,909 (34,291 males and 39,618 females) were enrolled. Most individuals were recruited from the Universities of Alberta, Toronto, and Western Ontario, but a small group was recruited through the Canadian Cancer Society. Participants completed baseline lifestyle and food frequency questionnaires and provided hair and toenail specimens.^9^ Cancer incidence was collected via linkage to the Canadian Cancer Registry. Registry diagnosis and histology were obtained via pathology reports and deaths were ascertained from linkage to the National Mortality Database. Follow-up was continued through December 2010 for those from Ontario, and through 2005 for residents of other provinces. For BCRPP, CSDLH provided a case-cohort including all 1,090 female breast cancer cases and a sub-cohort of 4,140 women aged 23-70 years.

The Janus Serum Bank Cohort (JANUS)

The Janus Serum Bank is a population-based biobank for cancer research in Norway. Blood serum samples were collected from 1972-2004 through various health examinations in Norway and from blood donors in Oslo, with a total of 318,628 Norwegians contributing.^10^ Five health examination studies were included, each with a different enrollment profile, covering men and women aged 20-42 years, while the blood donor donations included individuals 18-65 years. Average age at enrollment for the full cohort was 41 years. 90% of the cohort completed a baseline questionnaire with information on lifestyle factors, diet, anthropometrics, and other health markers.^10,11^ For BCRPP, missing baseline and follow-up information was obtained through a linkage to questionnaires sent to women invited to BreastScreen Norway during 2006-2015.^12^ Janus cohort members are followed from the time of their baseline questionnaire until emigration or death through linkage of participant members to the National Population Register and the Norwegian Cause of Death Registry. Information on cancer diagnoses is obtained through linkage to the Cancer Registry of Norway. All linkages used the unique personal identification numbers given to all legal residence since the 1960s. Additional details are found online: <https://www.fhi.no/en/cancer/janus-serumbank/> and <https://www.fhi.no/en/cancer/screening/breastscreen/epidemilogical-questionnaire-q2006/>.

Melbourne Collaborative Cohort Study (MCCS)

The MCCS was set up to investigate the role of diet and lifestyle factors in chronic diseases, with particular attention to cancer. 41,513 individuals (24,469 females and 17,044 males) aged 27-76 years (99% aged 40-69 years) were recruited from the Melbourne metropolitan area between 1990-1994, with deliberate oversampling of Southern European migrants (24% of the cohort). At baseline, individuals completed face-to-face interviews with diet and lifestyle questionnaires, physical measurements (e.g., fat mass), and food frequency questionnaires and gave a blood sample. Over time, incident cancers and deaths are identified by regular matching to cancer and death registries. Mailed questionnaires were sent 3-4 years after baseline and collected information on other health events (1995-1998). A second follow-up was completed 2003-2007, where participants were seen face-to-face in clinic for interviews and updated measurements.^13^ For BCRPP, data was shared from the subcohort that completed the second follow-up, at which time they were re-consented. This timepoint served as baseline for our study.

Mayo Mammography Health Study (MMHS)

The MMHS includes women without breast cancer aged 35 years and older living in Minnesota, Wisconsin, or Iowa who had screening mammography at the Mayo Clinic, Rochester, between 2003 and 2006. A total of 19,924 participants were enrolled and completed baseline questionnaires. Cancer incidence was followed via Mayo Clinic tumor registries and linkage to the state cancer registries.^14^ A case–cohort design was used to efficiently target mammogram collection to a subcohort of 2300 women plus all women with incident breast cancer. For BCRPP, this full case-cohort data was shared, including 1,364 breast and the subcohort members, aged 35-92 years. Additional study details are also provided on the NCI website: https://cedcd.nci.nih.gov/cohort?id=125.

Multiethnic Cohort Study (MEC)

Between 1993 and 1996, the Multiethnic Cohort Study recruited adults aged 45-75 living in California and Hawaii to investigate cancer risk factors and disease distributions across diverse populations. Cancer diagnoses are confirmed via state tumor registries. Baseline questionnaires collected demographics, medical and reproductive histories, medication use (including hormonal replacement therapy), family history of various cancers, smoking history, physical activity, and an extensive quantitative food frequency questionnaire.^15^ A total of 215,251 individuals were enrolled at baseline, including 111,186 women.^16^ A prospective biorepository was established between 2001-2006, yielding a subcohort of approximately 70,000 individuals to allow for genetic and other biomarker-based studies.^16–18^ Details can be found online: https://uhcancercenter.org/mec.

Nurses’ Health Study (NHS)

The NHS is a prospective cohort study started in 1976 enrolled 121,700 registered nurses aged 30-55 living in the 11 most populous states (California, Connecticut, Florida, Maryland, Massachusetts, Michigan, New Jersey, New York, Ohio, Pennsylvania, and Texas). The study was developed with the goal to evaluate risk factors for and risk of chronic diseases, including cancer.^19,20^ Every 2 years, participants complete detailed questionnaires including information on disease status, lifestyle, and reproductive factors. Over the years, a variety of biospecimens have been collected, including toenail samples (1982-1984), blood (1989-1990 and 2000-2002), and urine (2000-2002), and cheek cells (2001-2004). A food frequency questionnaire was added in 1980 and completed every 4 years. Participants are followed up for disease outcomes are self-reported or captured through linkage to cancer registries, (with confirmation of disease by medical records), and linkage to the National Death Index. See the website for additional cohort details: https://nurseshealthstudy.org/

Nurses’ Health Study 2 (NHS2)

NHS2 was started in 1989 and sought to explore oral contraceptive use, diet, and lifestyle risk factors and chronic diseases in a younger cohort of U.S. nurses (aged 25-42 at enrollment). A total of 116,430 women were enrolled across 14 states. Biennial questionnaires have been completed, and include information on disease status, lifestyle, and reproductive factors.^20^ Every 4 years, starting in 1991, participants complete detailed food frequency questionnaires. Blood and urine samples were collected from 30,000 individuals between 1996-1999 and a second sample was taken in 2010-2012. Cheek cells were collected in 2006. Follow-up occurs via self-report, disease registry linkages, and linkages to the National Death Index. Additional details are found online: https://nurseshealthstudy.org/

New York University Women’s Health Study (NYUWHS)

NYUWHS enrolled 14,274 women ages 35-65 years between 1985 and 1991. Recruitment was conducted through a mammography screening center in NYC. At enrollment, women completed detailed questionnaires and provided a blood sample. Participants completed up to six follow-up questionnaires, sent every 3-5 years to collect updated information on disease status, lifestyle and diet habits, and reproductive history.^21^ Cancer cases are followed via linkage to state registries. Additional details are online: https://med.nyu.edu/departments-institutes/population-health/divisions-sections-centers/epidemiology/nyu-womens-health-study.

The Prostate, Lung, Colorectal, Ovarian Cancer Screening Trial (PLCO)

The PLCO Cancer Screening Trial was a randomized controlled trial conducted by the National Cancer Institute to evaluate the efficacy of using screening tests to reduce cancer-related mortality from prostate, lung, colorectal, and ovarian cancer. Approximately 155,000 participants, aged 55-74 years, were enrolled from 1993-2001 from ten screening centers across the U.S. Information on cancer diagnoses, including non PLCO cancers, was abstracted from medical records. Deaths were ascertained via reports from relatives/friends/physicians, and linkage to the National Death Index. Blood and buccal cells were collected for research purposes and are stored at the Biorepository. The initial follow-up period was 13 years from randomization, though extended data collection began in 2009.^22,23^ A summary of the trial is provided in the NCI Cancer Data Access System: https://cdas.cancer.gov/learn/plco/trial-summary/

The Sister Study (SIS)

The Sister Study was started by the National Institute of Environmental Health Sciences in 2003 to evaluate how environmental and genetic factors influence risk of breast cancer. Between 2003 and 2009, the study enrolled 50,884 women 35-74 years of age with no prior breast cancer, who had a sister diagnosed with breast cancer, living in the US, including Puerto Rico. At baseline women completed detailed questionnaires to provide information on sociodemographics, family history of cancer, environmental and lifestyle exposures, employment, sleep habits, diet (via the Block 98 FFQ), and reproductive/pregnancy information. In addition, over 99% of the cohort provide blood, urine, toenail, and dust samples at baseline. Participants complete health update questionnaires each year to provide disease status. Every 2-3 years they answer detailed questionnaires to update data on risk factors as well. Cancer outcomes are noted on these updates and details are confirmed via medical record review.^24^ Additional details are found on the website: https://sisterstudy.niehs.nih.gov/english/index1.htm

Southern Community Cohort Study (SCCS)

SCCS was established in 2001 to better understand causes of cancer and chronic diseases. Between 2002-2009, 84,069 adults aged 40-79 years were enrolled in the study across the southeastern U.S. The majority of participants were recruited from community health centers specifically serving low-income, medically uninsured or underinsured individuals. Participants provided information on demographics, medical history, and lifestyle factors at baseline, as well as blood or urine specimens. To date, participants have been asked to update information on four rounds of follow-up questionnaires. Disease outcomes and deaths are tracked via linkage to registries.^25,26^ Information is also provided on their website: https://www.southerncommunitystudy.org/

United States Radiological Technologists Study (USRT)

USRT is a joint effort of the University of Minnesota, the National Cancer Institute Radiation Epidemiology Branch, and the American Registry of Radiologic Technologists started in 1982. The study began with the goal of investigating how low-dose occupational exposure to radiation relates to health outcomes. Certified radiologic technologists who were certified for at least 2 years between 1926-1982 were eligible; over 110,000 individuals have participated in the cohort, with baseline surveys completed between 1983-1989. The survey collected information on family history, risk factors, and radiation exposure. Three follow-up surveys have been completed (1994-1998, 2003-2005, 2012-2014) to update information and provide new information about nightshift work, physical activity, and other exposures.^27,28^ Details are found on the cohort website: https://dceg.cancer.gov/research/who-we-study/cohorts/us-radiologic-technologists

Women’s Health Initiative (WHI)

The WHI was initiated in 1992 to investigate prevention strategies for chronic diseases common to postmenopausal women. Women aged 50-79 years were enrolled in clinical trials on hormone therapy, diet modification, and/or calcium/vitamin D supplementation, between 1993-1998 at 40 US clinical centers. Those ineligible or uninterested in the trials were asked to participate in the observational study, and 161,808 women were enrolled.^29,30^ For the observational study, standardized questionnaires including demographic and risk factor information, alongside family and medical history were completed by participants. Physical measures (height, weight, blood pressure) were taken by certified staff at clinic visits and blood samples were also banked. Annual detailed follow-up surveys were completed for 8 years. Data collection was completed in 2005 for the original study, though extension studies have been done to collect long-term data from 52,068 volunteers, currently extending to 2026. Additional resources are found online: https://www.whi.org/

Women’s Health Study (WHS)

The WHS was a randomized trial started in 1993 to test low-dose aspirin and vitamin E to prevent heart disease and cancer in women.^31–34^ A total of 39,876 women aged 45 years or older in the U.S. were enrolled. At baseline, 71% of women provided a blood sample; they are the participants in the Women’s Genome Health Study.^32^ At baseline, 6-months, 12-months, and annually thereafter, women completed detailed questionnaires on compliance, illness/adverse effects, and risk factors. Participants self-reported non-fatal health outcomes on follow-up questionnaires; self-reports of cardiovascular disease and cancer were validated using medical records. Deaths were confirmed via medical records and/or death certificates, and the National Death Index was searched periodically. Dietary assessments were collected at baseline and in 2004. The trial ended as scheduled in 2004, following which 33,682 participants agreed to be followed observationally. Women in the follow-up cohort are asked to complete detailed follow-up questionnaires on medical history and lifestyle risk factors on an annual basis. Details can be found on the website: https://whs.bwh.harvard.edu/index.html.

**Data harmonization for BCRPP cohorts**

To date, the following data has been shared on female cohort members: (a) baseline participant characteristics (including demographics, anthropometrics, reproductive factors, personal history of benign breast disease or biopsies, family history of breast and ovarian cancer, and behaviors such as smoking, alcohol use, and physical activity), (b) details on first and second incident breast cancer diagnoses (including year and age of diagnosis, stage, grade, tumor size, ER/PR/HER2 status, and laterality), and (c) mammographic density.

Cohorts were asked to create a BCRPP-specific dataset that aligned with our detailed data dictionary. To avoid issues in data harmonization from the outset, cohorts performed quality control checks (code provided by BCRPP) and BCRPP members requested changes from individual cohorts based on outputs before receiving final datasets. Where variables were not collected in a way that matched the formatting of the BCRPP data dictionary, cohorts provided ancillary files that were reviewed by BCRPP members and harmonized within the data dictionary. For instance, if a cohort provided smoking amount in categorical data in ancillary files, individuals were assigned the median of their respective category to match other cohorts. Extensive data checks were performed after receiving finalized cohort-specific datasets, and further harmonization was performed where required. The complete data dictionary for this study can be accessed via the BCRPP data platform: <https://epidataplatforms.cancer.gov/bcrpp>.

Future data collection will focus on obtaining follow-up data for cohorts with multiple questionnaire cycles, as well as details on genetic risk factors, including polygenic risk scores (PRS).

**Revision of iCARE-calibrate function**

To test model performance, we used a modified version of the iCARE-calibrate function **(**iCARE package, v.1.36.0 available via BiocManager). Base code is available publicly on GitHub ([parichoy/iCARE](https://github.com/parichoy/iCARE/tree/devel)). Modifications included different handling of ties for AUC calculation and censoring at last follow-up time. For the AUC calculation, we handled ties by assigning 0.5 for each pair where predicted risk in control=predicted risk in cases. For censoring, we applied a change to the code wherein risk was only calculable from the timepoint 0 to the minimum of either 5 years or the maximum follow-up time for individuals, as not all individuals had a full 5-years of follow-up.

**Modeling assumptions**

Some variables that are part of the risk factor models used were not collected or were unable to be harmonized and were set to missing in risk prediction. These included history of prostate cancer and bilateral oophorectomy (BWHS RC) and details on BC or ovarian cancer from second- or third-degree relatives (TC, BOADICEA).

We used the provided first-degree family history information along with several assumptions to allow for risk calculation using TC and BOADICEA. Family history information collected in BCRPP included: first-degree family history of breast or ovarian cancer (yes/no), number of first-degree relatives with breast or ovarian cancer, number of relatives with BC diagnosed before age 50, whether the mother or father had breast cancer, number of sisters, number of daughters, and the number of sisters and daughters that had breast cancer. Among studies with a reported first-degree family member with BC, but no specification as to which relative was diagnosed, we assumed the individual’s mother was affected, with diagnosis age set as missing if no age was provided, 70 y if diagnosis age was given as >50 y and 45 y if diagnosis age was given as <50 y. These ages were selected based on SEER averages for BC diagnosed under 50 y or over 50 y of age (SEER Incidence Data, November 2025 Submission (1975-2023)). In sensitivity analyses we assumed that first-degree family history of breast cancer was the individual’s sister or grandmother instead of their mother; however, here we present the first assumption given non-substantial differences in model performance.

In the TC model, if individuals reported a sister with breast cancer, we assumed the sister was diagnosed at the same current age as the proband. For the BOADICEA model, age and birth year was also required for each parent and sister, regardless of breast cancer status. Here we assumed parental age was equal to the proband’s age plus 30 years and sister’s age was the proband’s age plus 5 years. Affected sisters were assumed to be diagnosed at the proband’s current age to maintain consistency with the TC model.

**Reference Populations and Incidence Rates for iCARE-Lit**

The iCARE-Lit model allows for data imputation via a reference population that is intended to closely resemble the risk factor profile of the underlying population for the cohort of interest. For U.S.-based cohorts and CSDLH, we used the same reference population, with risk factor distributions derived from the National Health and Nutrition Examination Survey (NHANES) from 2008, 2010, and 2012, aside from alcohol, which was based on the distribution among controls from the Women’s Health Initiative (WHI) study. Details on the reference dataset creation and imputation within iCARE-Lit are provided elsewhere.^35^ Incidence rates for U.S. cohorts were taken from non-Hispanic White women in the Surveillance Epidemiology and End Results (SEER) (2000-2009); this was used for simplification purposes as most U.S. cohorts were comprised of non-Hispanic White women. For BGS, the reference population for imputation was created using individuals included in BGS, as well as population-level surveys. Incidence rates were applied from Cancer U.K. (2000-2016). For JANUS, the reference population consisted of individuals with complete-case data from the cohort, and Norwegian population-based BC incidence rates (2022) were applied.
