## Supplemental Figures for "Performance of general-population breast cancer risk prediction models in an international consortium"

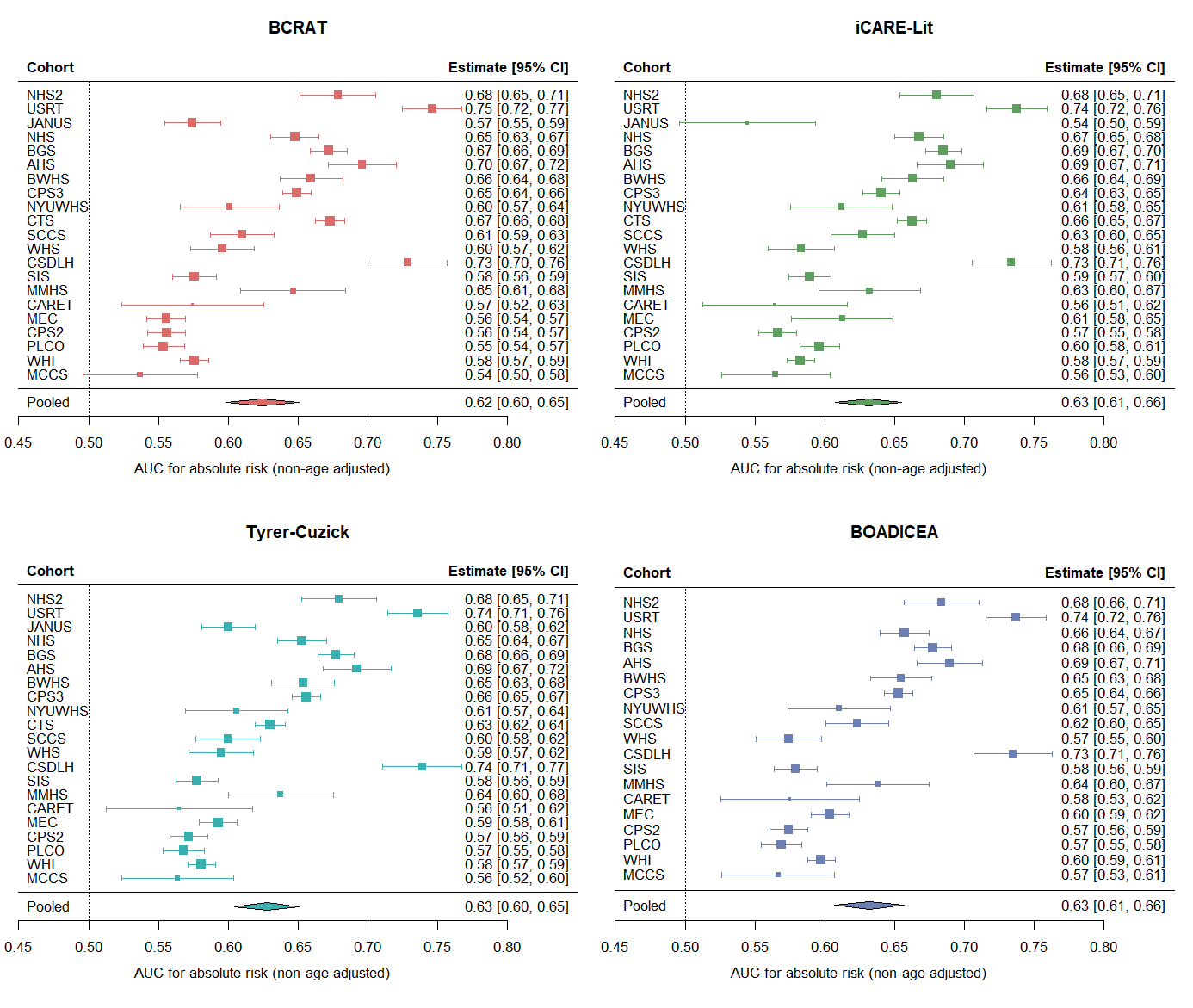

**Figure S1**. Overall AUC without age-adjustment based on absolute risk by model for all cohorts. Pooled estimates are given from random effects meta-analysis. Cohorts appear in order of mean age at baseline questionnaire (youngest to oldest from top to bottom). BCRAT: Breast Cancer Risk Assessment Tool; BOADICEA: The Breast and Ovarian Analysis of Disease Incidence and Carrier Estimation Algorithm; iCARE-Lit: Individualized Coherent Risk Estimation (iCARE) Literature Model. Note BOADICEA was not calculated for JANUS and CTS. I^2^ (total heterogeneity/total variability) BCRAT: 97.8%, iCARE-Lit: 97.1%, Tyrer-Cuzick: 97.4%, BOADICEA: 97.1%. P-heterogeneity <0.001 for all models. Note, the estimate for iCARE-Lit for JANUS included only postmenopausal women (N=12,333) as there were a limited participants with complete information on covariates among premenopausal women, precluding creation of a premenopausal reference dataset for this cohort.

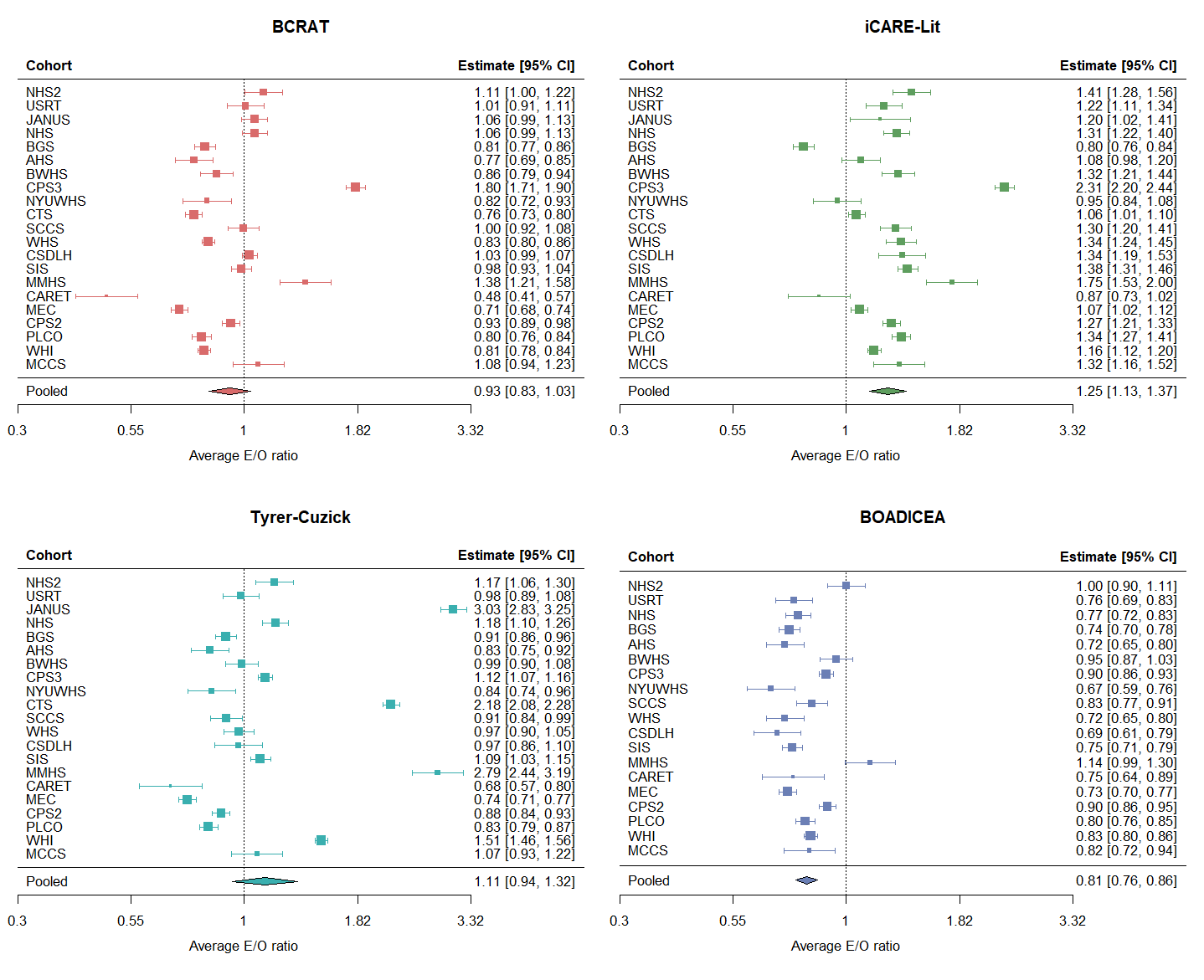

**Figure S2.** Expected to observed (E/O) ratios on average by cohort and model. Cohorts are listed by age at baseline from lowest to highest. E/O ratios are shown on the log scale.

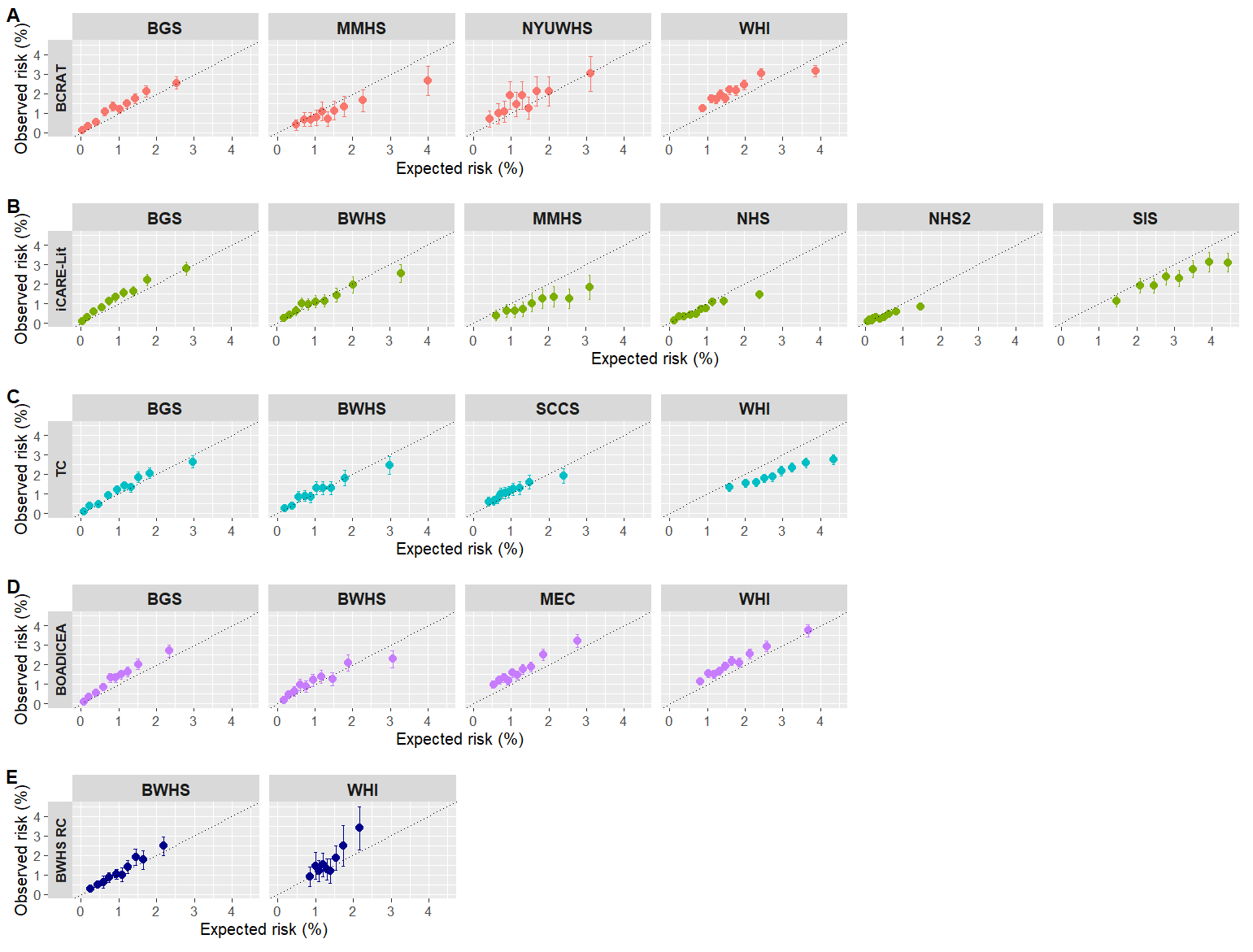

**Figure S3**. Observed (95% CI) vs. expected absolute risk by cohort for those with all required model variables for (a) BCRAT, (b) iCARE-Lit, (c) TC, (d) BOADICEA, (e) BWHS. Observed and predicted values and observed confidence intervals were truncated at 5% for visualization purposes. BCRAT: Breast Cancer Risk Assessment Tool; BOADICEA: The Breast and Ovarian Analysis of Disease Incidence and Carrier Estimation Algorithm; iCARE-Lit: Individualized Coherent Risk Estimation (iCARE) Literature Model; TC: Tyrer-Cuzick mode. For TC and BOADICEA, cohorts considered to have complete data if they included all requested family history variables in BCRPP (see Supplemental Methods for additional assumptions).

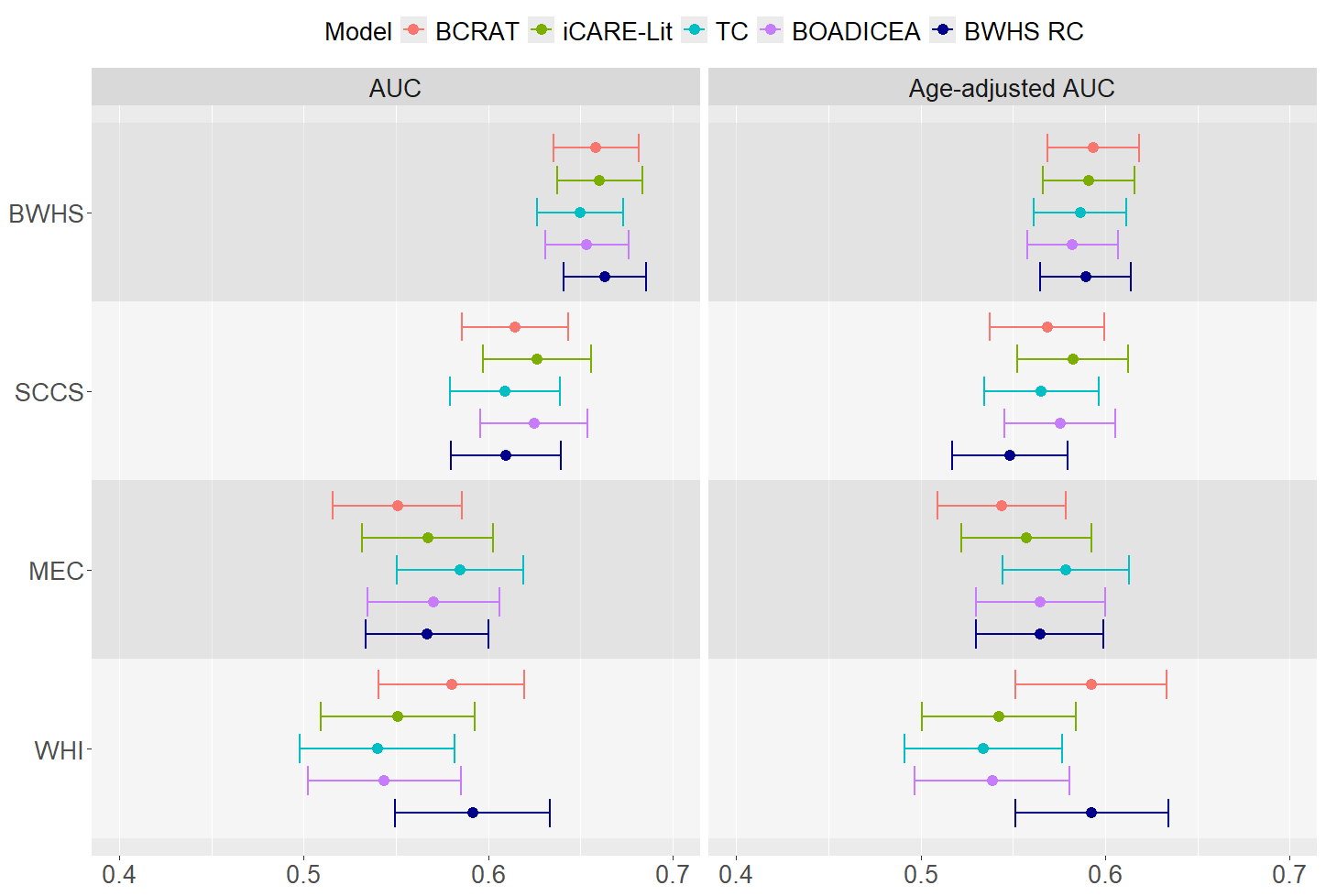

**Figure S4**. AUC and age adjusted AUC comparisons among Black women aged 30-70 years by model. Cohorts represent those with at least 100 invasive BC cases in 5 years.
