## Supplementary material for "Performance of general-population breast cancer risk prediction models in an international consortium": Funding information and author contributions

**Cohort Funding and Acknowledgements**

Agricultural Health Study (AHS)

The AHS was supported by the National Institute of Environmental Health Sciences Z01 ES049030 and National Cancer Institute grants ZIACP010119 and ZIACP010123.

Breast Cancer Now Generations Study (BCNGS)

Breast Cancer Now, the UK National Health Service, and the Institute of Cancer Research.

Black Women’s Health Study (BWHS)

US National Institutes of Health (NIH; U01-CA164974). Dr. Palmer’s research is supported by National Institutes of Health grant R01CA164974, the Susan G. Komen Foundation, the Breast Cancer Research Foundation, and the Karin Grunebaum Cancer Research Foundation.

The authors would like to acknowledge the contribution to this study from central cancer registries supported through the Centers for Disease Control and Prevention’s National Program of Cancer Registries (NPCR) and/or the National Cancer Institute’s Surveillance, Epidemiology, and End Results (SEER) Program. Central registries may also be supported by state agencies, universities, and cancer centers. Participating central cancer registries include the following: AL, AR, AZ, CA, CO, CT, DE, DC, FL, GA, HI, IA, IL, IN, KY, LA, MD, MA, MI, MO, MS, NE, NJ, NM, NY, NC, OH, OK, OR, PA, SC, TN, TX, VA, SEER Seattle, WI. The content is solely the responsibility of the authors and does not necessarily represent the official views of the U.S. Department of Health and Human Services, the National Institutes of Health, the National Cancer Institute, or the state cancer registries. We thank participants and staff of the BWHS for their contributions.

Carotene and Retinol Efficacy Trial (CARET)

Supported by grants (U01-CA63673, UM1-CA167462, and U01-CA167462) from the National Cancer Institute.

California Teachers Study (CTS)

NIH National Cancer Institute (NCI; award numbers U01-CA199277, P30-CA033572, P30-CA023100, UM1-CA164917, and R01-CA077398), cancer incidence data was funded by the California Department of Public Health pursuant to California Health and Safety Code (section 103885); US Centers for Disease Control and Prevention (CDC) National Program of Cancer Registries (NPCR; cooperative agreement 5NU58DP006344); the NCI Surveillance, Epidemiology and End Results (SEER) Program (contract HHSN261201800032I [awarded to the University of California, San Francisco, CA, USA], contract HHSN261201800015I [awarded to the University of Southern California, Los Angeles, CA, USA], and contract HHSN261201800009I [awarded to the Public Health Institute]).

Cancer Prevention Study 2 & 3 (CPS2 & CPS3)

The American Cancer Society funds the creation, maintenance, and updating of the Cancer Prevention Study-II and Cancer Prevention Study-3 cohorts. The authors express sincere appreciation to all Cancer Prevention Study-II and Cancer Prevention Study-3 participants, and to each member of the study and biospecimen management group. The authors would like to acknowledge the contribution to this study from central cancer registries supported through the Centers for Disease Control and Prevention's National Program of Cancer Registries and cancer registries supported by the National Cancer Institute's Surveillance Epidemiology and End Results Program.

Canadian Study of Diet, Lifestyle, and Health (CSDLH)

Establishment of the cohort was supported by grants from the National Cancer Institute of Canada and from the Canadian Institutes of Health Research. TER is supported by the Breast Cancer Research Foundation (BCRF-24–140).

The Janus Serum Bank Cohort (JANUS)

The Janus Serum Bank Cohort was formally established in 1973 and was financed by the Norwegian Cancer Society until 2004. After 2004, the biobank was transferred to the Cancer Registry of Norway, which currently manages the material and is responsible for the administration and operation of the biobank.

Melbourne Collaborative Cohort Study (MCCS)

Cancer Council Victoria, VicHealth and the Australia National Health and Medical Research Council (209057, 396414, 1074383)

Multiethnic Cohort Study (MEC)

The MEC is supported by the National Cancer Institute grant U01CA164973.

Mayo Mammography Health Study (MMHS)

The cohort was supported by NIH (grants R01CA140286, R01CA097396) and Mayo Clinic Cancer Center.

Nurses’ Health Study (NHS)

National Institutes of Health grants UM1CA186107, P01CA87969, R01CA49449. The content is solely the responsibility of the authors and does not necessarily represent the official views of the National Institutes of Health.

Nurses’ Health Study 2 (NHS2)

National Institutes of Health grants R01CA50385, U01CA176726, and U01HL145386. The content is solely the responsibility of the authors and does not necessarily represent the official views of the National Institutes of Health.

New York University Women’s Health Study (NYUWHS)

NIH U01CA290680, R01CA098661, UM1CA182934 and center grants P30CA016087 and P30ES000260

The Prostate, Lung, Colorectal, Ovarian Cancer Screening Trial (PLCO)

The NIH Intramural Research Program grants ZIACP010207, ZIACP010152

The Sister Study (SIS)

The National Institute of Environmental Health Sciences (NIEHS) Intramural Research Program (Z01-ES044005)

Southern Community Cohort Study (SCCS)

The SCCS is supported by the National Cancer Institute (grants R01 CA092447 and U01 CA202979) and supplemental funding from the American Recovery and Reinvestment Act (3R01 CA 029447-0851).

United States Radiological Technologists Study (USRT)

This research was supported by the Intramural Research Program of the National Institutes of Health (NIH) (ZIACP010133).

Women’s Health Initiative (WHI)

The WHI program is funded by the National Heart, Lung, and Blood Institute, National Institutes of Health, U.S. Department of Health and Human Services through contracts 75N92021D00001, 75N92021D00002, 75N92021D00003, 75N92021D00004, 75N92021D00005.

Women’s Health Study (WHS)

The WHS is supported by grants CA047988, HL043851, HL080467, HL099355, and UM1 CA182913 from the National Institutes of Health.

Note: The contributions of the NIH authors were made as part of their official duties as NIH federal employees, are in compliance with agency policy requirements, and are considered Works of the United States Government. However, the findings and conclusions presented in this paper are those of the authors and do not necessarily reflect the views of the NIH or the U.S. Department of Health and Human Services.

**Author Contributions**

All authors critically reviewed the article and provided final approval of the version to be published and are accountable for all aspects of the work. Additional contributions are below:

KDB: Acquisition of data, data harmonization, formal analysis, interpretation of data, writing of the original draft, conception and design

TUA: Acquisition of data, interpretation of data, project oversight, conception and design

ELN: Data harmonization, interpretation of data, writing of the original draft

ABG: Acquisition of data

RM: Acquisition of data, interpretation of data, writing of the original draft

JP: Acquisition of data, interpretation of data, writing of the original draft

RTF: Acquisition of data, interpretation of data, writing of the original draft

CMV: Acquisition of data, interpretation of data, writing of the original draft

LBF: Acquisition of data

RF: Acquisition of data, data harmonization, formal analysis, interpretation of data

KAB: Acquisition of data

GZ: Acquisition of data, data harmonization, interpretation of data

MLN: Acquisition of data

MB: Acquisition of data, data harmonization

LRT: Acquisition of data

JMH: Acquisition of data, data harmonization

AVP: Acquisition of data

CB: Acquisition of data

JVL: Acquisition of data

ESS: Acquisition of data, data harmonization, formal analysis, interpretation of data

TER: Acquisition of data

VAK: Acquisition of data

HL: Acquisition of data

KMT: Acquisition of data, data harmonization, formal analysis, interpretation of data

RLM: Acquisition of data, data harmonization

CH: Acquisition of data

CGS: Acquisition of data, data harmonization

AHE: Acquisition of data

BR: Acquisition of data

WCW: Acquisition of data

ARN: Acquisition of data

YC: Acquisition of data

FW: Acquisition of data, data harmonization

WZ: Acquisition of data

JL: Acquisition of data, data harmonization

KMO: Acquisition of data, data harmonization

DPS: Acquisition of data

CMK: Acquisition of data, data harmonization

MSL: Acquisition of data

GA: Acquisition of data

JCL: Acquisition of data, data harmonization

IML: Acquisition of data

MCGC: Conception and design, interpretation of data, project oversight

NC: Conception and design, interpretation of data, project oversight

PK: Conception and design, interpretation of data, project oversight
